# Demonstrating the Beneficial Effect of Low Protein Diet in Primary Sclerosing Cholangitis through a Randomized Clinical Trial and Multi-omics Data Analysis

**DOI:** 10.1101/2024.02.23.24303167

**Authors:** Xiaole Yin, Gila Sasson, Zheng Sun, Shanlin Ke, Demsina Babazadeh, Shaikh Danish Mahmood, Macie Andrews, Shelley Hurwitz, Tinashe Chikowore, Maia Paul, Nadine Javier, Malav Dave, Alexandra Austin, Linda Gray, Francene Steinberg, Elaine Souza, Christopher Bowlus, Yang-Yu Liu, Joshua Korzenik

## Abstract

Primary sclerosing cholangitis (PSC), a progressive cholestatic hepatobiliary disease characterized by inflammation and fibrosis of the bile ducts, has a pathophysiology that is not understood. No effective therapies exist. The only treatment option for PSC is liver transplant. We undertook a pilot randomized trial of diet to investigate the pathophysiology of the disease, the role of diet and to advance potential therapy. We enrolled 20 patients with PSC and randomly assigned them to a Low Protein/low sulfur Diet (LPD, n=10) or the Specific Carbohydrate Diet (SCD, n=10) for 8 weeks. Results showed that low protein intake benefits PSC patients, whereas higher protein levels exacerbate the condition. We further identified gut bacterial markers useful for distinguishing LPD responders (mostly PSC with concomitant ulcerative colitis) from non-responders. Additionally, by integrating multi-omics data, we propose that this diet modifies the intestinal sulfur cycle reducing hydrogen sulfide (H_2_S) production. Our findings provide an understanding of the beneficial effect of LPD as well as insights into a possible key driver of inflammation in PSC.

## INTRODUCTION

Primary sclerosing cholangitis (PSC), a chronic cholestatic liver disease characterized by inflammation and fibrosis of the biliary tree, can lead to debilitating consequences including end-stage liver disease and malignancies. Currently, no therapy has been found to be effective for PSC, other than liver transplant, which remains the mainstay of therapy. However, the disease can recur after transplant. PSC is intimately associated with inflammatory bowel disease (IBD), particularly ulcerative colitis (UC). Approximately 70-80% of patients with PSC also have underlying IBD [1, 2], but only around 3-5% of UC patients will develop PSC [3]. Thus, the combination of the two creates a distinct disease phenotype, which is a prototypical disease of the gut-liver axis [1, 4].

The pathophysiology of PSC remains poorly understood. The paradigm of IBD is relied on as a model to understand PSC, presuming an interaction between the environment, the microbiome and immune system with a genetic predisposition [5, 6]. Other hypotheses suggest a toxic bile acid or a cross reacting T cell response between the colonocytes and cholangiocytes [5, 7]. The immune modifying approaches that can be effective in IBD do not benefit PSC. A distinct dysbiotic fecal microbiome has been detailed in PSC, which overlaps with that of IBD [1, 4, 8]. While numerous studies have investigated changes in the gut microbiome in the context of IBD [9–12], fewer have focused on PSC [13–18].

Our approach to PSC in this study is based on the hypothesis that sulfur-containing amino acids such as methionine may be substrates for sulfate-reducing bacteria to generate hydrogen sulfide (H_2_S). H_2_S production is likely coupled with increased bacterial degradation of mucin, which is rich in cysteine and methionine [19, 20]. Further, H_2_S can also impair colonocyte uptake and beta-oxidation of butyrate [21]. At higher levels, H_2_S can be toxic to mitochondria, generating radical oxygen species and activating the NLRP10 inflammasome, generating a broader inflammatory reaction [22]. In this way, reducing H_2_S by manipulating dietary sulfur content may limit cellular impairment and inflammation. Practically speaking, this would involve the reduction of sulfur-containing amino acids, which are primarily derived from animal and plant-based proteins.

In contrast, the comparator diet examined in this study, the Specific Carbohydrate Diet (SCD), is a popular diet among IBD patients that is grain-free and low in sugar and lactose, but does not limit protein content. This diet has been studied to a limited degree in IBD with variable effectiveness [23, 24]. The SCD excludes the intake of complex carbohydrates, disaccharide-containing foods, grain, corn, potato, and dairy while permitting carbohydrates obtained from fruits, honey, some vegetables, and fermented yogurt. The SCD is based on the principle that specifically selected carbohydrates in SCD require minimal digestion, so they are subsequently well absorbed, avoiding further gut microbial metabolism.

Diet composition may influence gut microbial activity in PSC, both through supporting or enhancing the growth of particular bacterial communities and modifying metabolic profiles. These adaptations may consequently influence disease progression through the gut-liver axis [25–29]. Yet, the interplay between these parameters is complex and poorly understood [4] especially with regard to its effect on disease course [30, 31]. Despite the strong patient interest, clinical trials targeting dietary interventions specifically for PSC are notably lacking. To date, only one study has examined a gluten-free diet in individuals with PSC and associated colitis, and it failed to demonstrate clinical or biomedical improvement [32].

In this study, we conducted a pilot randomized controlled trial to assess the clinical efficacy of a low-protein diet (LPD) compared to the SCD in PSC (**Fig. 1a & 1b**), and leveraged high-throughput technologies (shotgun metagenomic sequencing and global untargeted metabolomic profiling) and multi-omics data analyses to gain mechanistic insights into response to dietary intervention (**Fig. 1c**).

**Figure 1.**
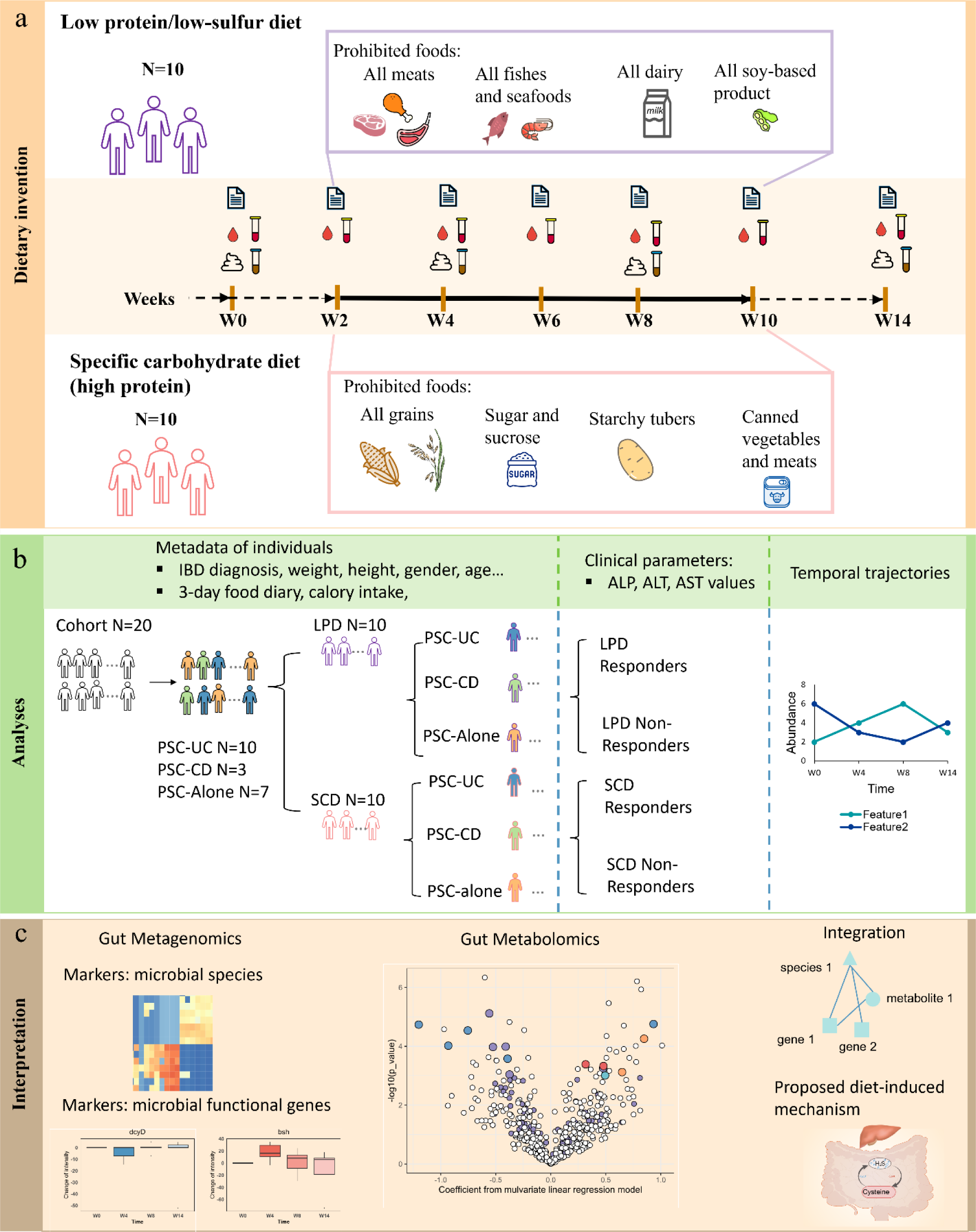
Overview of the study design. Two dietary interventions: Low protein/low sulfur diet (LPD) and Specific carbohydrate diet (SCD) were compared. We profiled fecal metagenomic and metabolomic data at four time points. 3-day dietary records, co-diagnosis of IBD (UC, CD, or non-IBD), and clinical parameters were collected. After dietary interventions, the response outcomes and temporal trajectory were analyzed and interpreted through multiple perspectives.

## RESULTS

### Patient characteristics

We screened ∼100 individuals with PSC, and 25 met eligibility criteria and 5 eligible individuals chose not to participate prior to randomization. Ten participants were randomly assigned to the LPD and ten to the SCD (**Fig. 1a**, **Fig. 1b & Table 1**). Participant characteristics were balanced between the groups. Two participants withdrew due to cholangitis, and their data until withdrawal were incorporated into the analysis.

**Table 1.** Patients’ characteristics at inclusion.

| **Table 1 Patients' characteristics at inclusion.** |  |  |
| --- | --- | --- |
|  | LPD (n=10) | SCD (n=10) |
| Age at inclusion (years) <sup>a</sup> | 43.79 ± 10.05 | 43.95 ± 8.99 |
| Gender (male/female) | 10 (7/3) | 10 (4/6) |
| Diagnosis <sup>b</sup> |  |  |
| PSC-UC | 6 (60%) | 4 (40%) |
| PSC-CD | 1 (10%) | 2 (20%) |
| PSC-alone | 3 (30%) | 4 (40%) |
| BMI <sup>a</sup> | 26.89 ± 6.69 | 23.53 ± 3.77 |
| Smoker=yes <sup>b</sup> | 3 (30%) | 2 (20%) |

<sup>a</sup> Mean ± SD
<sup>b</sup> n (%)
UC, ulcerative colitis; CD, Crohn's disease; IBD, inflammatory bowel disease; PSC, primary sclerosing cholangitis; BMI, Body mass index.

Participants in the LPD group demonstrated lower protein consumption in 3-day food recordings at all study time points during dietary intervention (week 2 to week 10) compared to SCD group participants (Wilcoxon rank-sum test, 4.57e-05 < p-value < 0.010), supporting the compliance with the dietary interventions (**Fig. 2a**).

**Figure 2.**
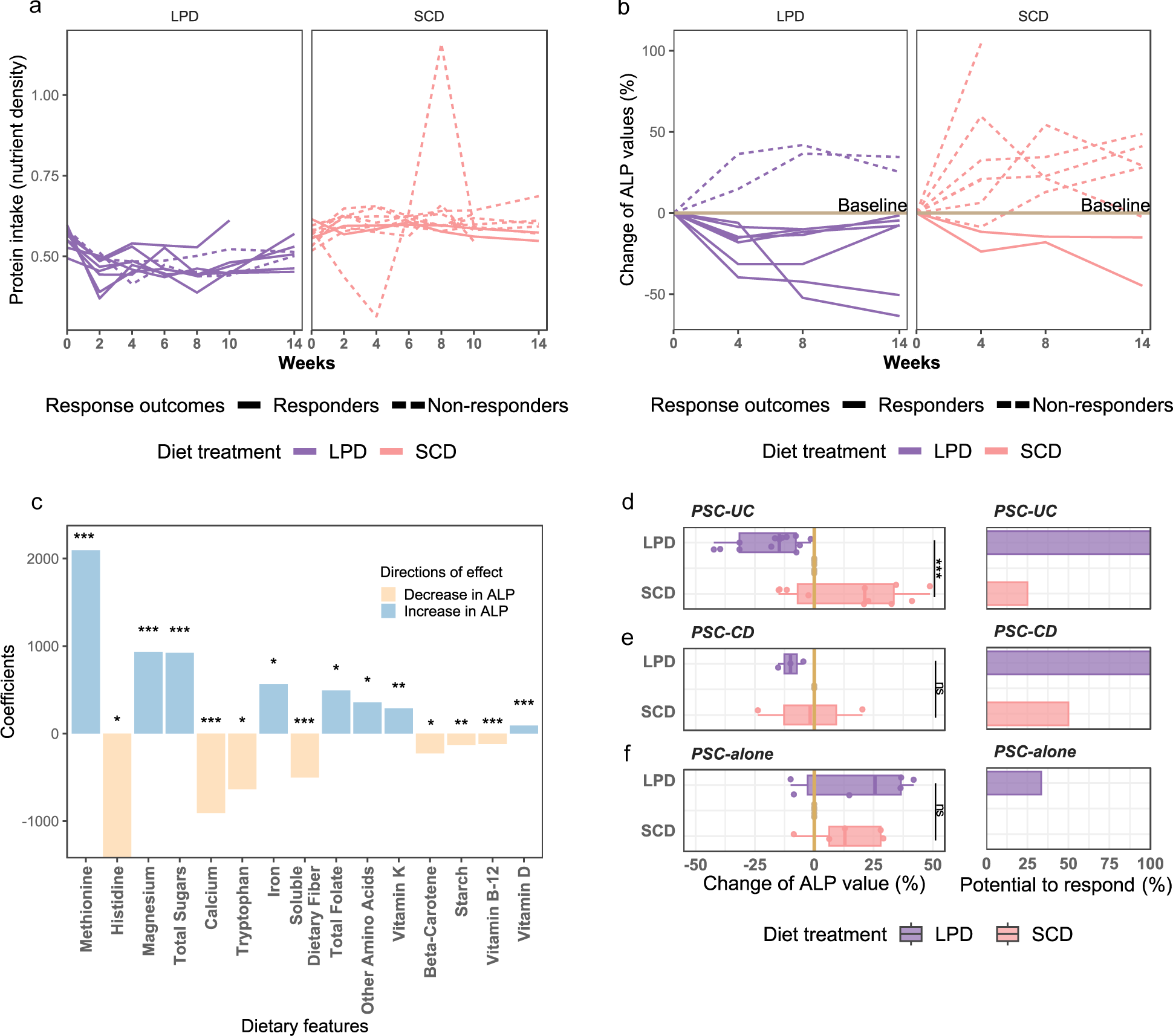
LPD dietary intervention benefits patients with PSC, whereas SCD exacerbates the condition. **a**) Protein intake in LPD and SCD. Each line represents an individual. Protein intake amount was adjusted by the total energy intake to estimate the nutrient density. **b**) Percent change in alkaline phosphatase (ALP) from baseline before, during and after dietary intervention. Each line represents an individual. **c**) Coefficients of selected nutrient features after Lasso regression, indicating that reduced methionine intake significantly lowers ALP values. Nutrients with significance in the regression model were plotted, with *** denoting strong significance (p-value < 0.001), ** very significant (0.001< p-value < 0.01), * significant (0.01< p-value < 0.05). **d-f**) Variations in ALP values and the potential rates to respond indicated differing responses among three disease categories, **d**) PSC-UC, e) PSC-CD, and f) PSC-alone. The Wilcoxon rank-sum test was employed to assess significance between LPD and SCD, with *** denoting strong significance (p-value < 0.001), and ‘ns’ indicating no significant difference (p-value > 0.05).

### LPD benefits PSC patients, but SCD does not

Comparing the ALP levels at W4 and W8 with those at W0, we found that LPD led to improvement (i.e., decrease in ALP) in eight of the ten individuals (80%), with six of them (75%) returning to their baseline ALP level when dietary interventions were removed at W10 (**Fig. 2b)**. In contrast, only two of the ten participants (20%) in the SCD group responded to SCD (i.e., with lower ALP levels than their baseline levels). Moreover, for most of the participants in the SCD group, their ALP values did not appear to be affected after discontinuing the dietary intervention, suggesting that SCD may not be a potentially effective treatment for PSC. For those eight participants who responded to LPD, hereafter referred to as LPD-responders, their ALP values were reduced by 18.5% ± 10.4% at W4 (i.e., after two weeks of treatment), and by 22.6% ± 15.6% at W8 (i.e., after another four weeks of treatment).

When accounting for an ALP decrease of at least 10%, LPD showed efficacy in 60% of patients, compared to a 20% efficacy rate in SCD. With a reduction of at least 20%, LPD demonstrated efficacy in 30% of patients, while SCD showed a 10% rate. Considering a decrease of at least 30%, LPD maintained its efficacy in 30% of patients, with no cases of efficacy in SCD. Finally, with an ALP decrease of at least 50%, LPD is effective in 10% of patients, with SCD also showing no cases of effectiveness. Changes of other parameters, including Aspartate Aminotransferase (AST) and Alanine Aminotransferase (ALT) values, during the intervention were shown in **Fig. S1**.

Direct evidence obtained from Lasso Regression analysis revealed a significant positive correlation between the dietary methionine levels and the ALP levels (**Fig. 2c)**. This finding indicates that lower methionine intake is associated with lower ALP levels compared to baseline values. However, it is crucial to note that this relationship is correlative rather than causative.

In addition to methionine, levels of magnesium, total sugar, iron, total folate, other amino acids, vitamin K and vitamin D were also positively associated with the ALP levels (**Fig. 2c)**. By contrast, the levels of histidine, calcium, tryptophan, soluble dietary fiber, beta-carotene, starch, and vitamin B-12 were negatively associated with the ALP levels. At the same time, we observed that carbohydrate consumptions (nutrient density) exhibited significant differences between LPD and SCD, with LPD being higher than SCD at all time points during dietary intervention (week 2 to week 10) compared to baseline (Wilcoxon rank-sum test, 0.0014 < p-value < 2.17e-05) (**Fig. S2**).

### Three disease categories respond differently to dietary intervention

We performed a stratified analysis to determine if comorbid IBD influences ALP response to dietary interventions. Though the numbers of individuals in each subgroup were small, marked differences were evident for certain disease categories. For instance, for PSC-UC patients (n= 10 in total), LPD (n=6) was significantly more effective in lowering ALP than SCD (n=4), (Wilcoxon rank-sum test, p-value = 0.038). Moreover, all the PSC-UC patients in the LPD arm responded to LPD assessed by decreasing ALP levels, see **Fig. 2d**. For PSC-CD patients (n= 3 in total), there was only one patient in the LPD arm (n=1), and this patient responded to LPD. Interestingly, for PSC patients without IBD (PSC-alone) (n=7 in total), one patient in the LPD arm (n=3) responded, while no response in SCD (n=4). Moreover, the two interventions didn’t demonstrate any significant difference in reducing ALP levels (**Fig. 2f**).

### Gut microbial features distinguish LPD responders from non-responders

Given the efficacy of LPD, we aimed to determine distinguishing microbial biomarkers between LPD responders and non-responders using shotgun metagenomic sequencing data. We found that the alpha diversity (measured by the Shannon index) of stool samples in the PSC-UC group was significantly lower than that in the PSC-CD and PSC-alone groups (**Fig. S3a & Fig. S3b,** Wilcoxon rank-sum test, p-value = 0.016 and p-value = 0.00062, respectively). Samples in each category were clustered together and distinct from the other two groups (PERMANOVA p-value=0.001, F=3.53) (**Fig. S2c**). When individuals were simply classified into responders (individuals showing reduced ALP) and non-responders (individuals showing reduced ALP), variations in alpha diversities and differences in beta diversity can be found in **Fig 3a, Fig. S4 & Fig. S5**. The clusters of LPD were separated from LPD non-responders in the PCoA plot based on the Bray-Curtis dissimilarity, suggesting that gut microbiota is key in differentiating response outcomes (**Fig. 3a**). Similar results were revealed by UMAP clustering (**Fig. S4b**).

**Figure 3.**
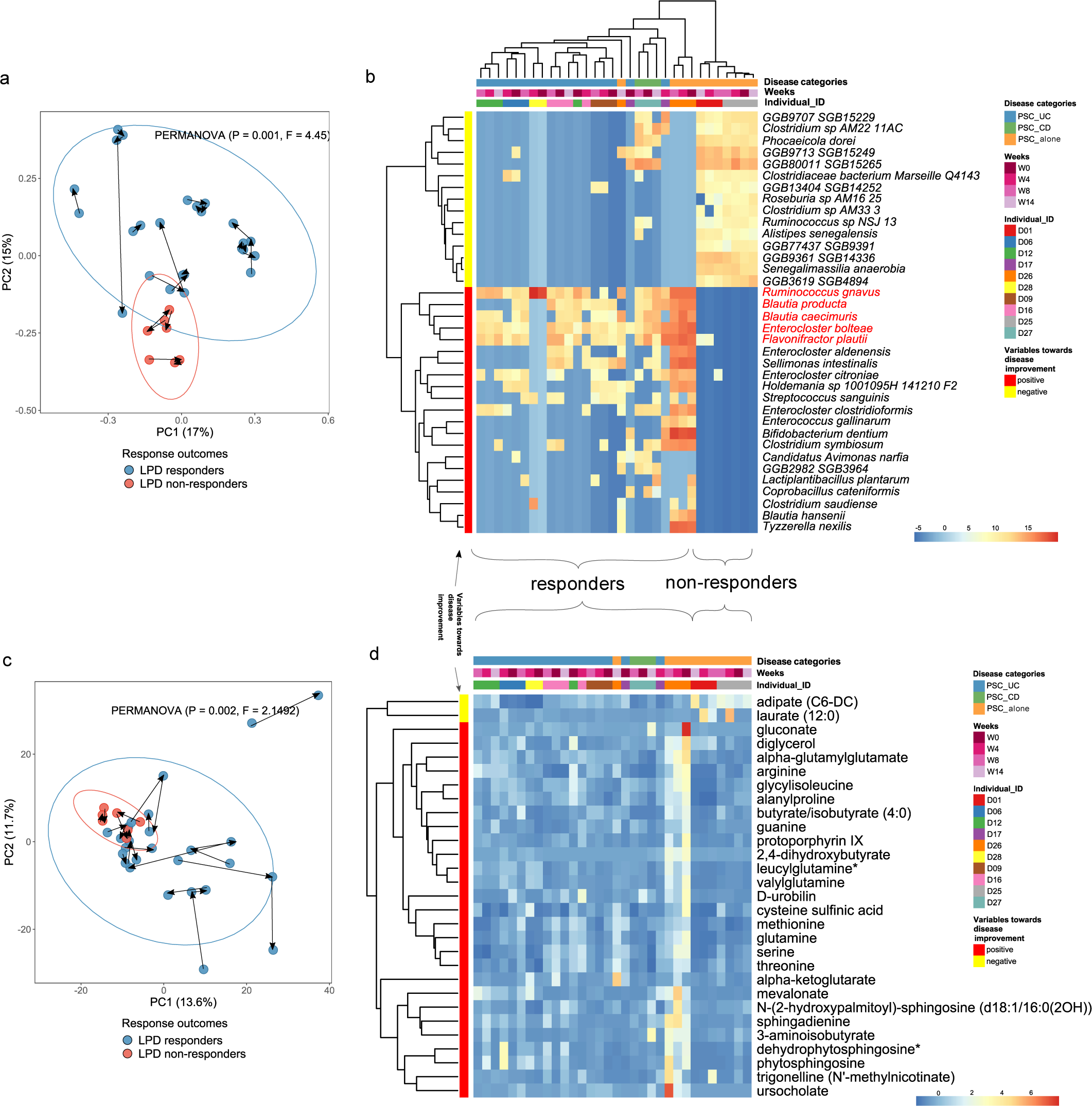
Fecal metagenomics predicts response outcomes. **a**) PCoA plot of the gut microbial compositions of LPD samples, differentiated by response outcomes (blue for responders: individuals showing decreased ALP, and pink for non-responders: individuals showing increased ALP, shows that LPD responders have distinct microbial structure from LPD non-responders. Arrows were drawn to connect samples from the same individuals in chronological order. **b**) Heatmap of bacterial species markers that can distinguish LPD responders from non-responders and the abundance distribution of these markers correlate with disease categories, as determined by multivariate linear regression models (target p-value < 0.05 and FDR < 0.2). Each column in the heatmap is a sample, and all the samples including both LPD responders and LPD non-responders are clustered based on the similarity of the bacterial composition. Each row is a species, and species are clustered based on the similarity of the distribution pattern across samples of the specified species. **c**) PCA plot of the gut metabolite compositions of LPD samples, differentiated by response outcomes (blue and pink), with arrows to connect samples from the same individuals in chronological order. **d**) Heatmap of metabolite markers that can distinguish LPD responders from non-responders. The columns in this heatmap are arranged to correspond directly with the order of the columns in the microbial profile heatmap in Fig. 3b.

To account for three disease categories as covariates and to eliminate random effects of inter-individual variation due to temporal changes, we employed MaAsLin2 to perform a multivariate linear regression analysis on the taxonomic profiles of the stool samples (see **Methods, Formula 1)**. We aimed to find microbial features that significantly differentiate LPD responders from LPD non-responders (with cutoff p-value < 0.05 and q-value < 0.2). We identified 21 (or 15) microbial species as significantly positively (or negatively) associated with responders, respectively (**Fig. 3b**). Among these, *Ruminococcus gnavus, Blautia caecimuris, Blautia producta, Enterocloster bolteae,* and *Flavonifractor plautii,* were more abundant in responders across samples of all three disease categories. Conversely, *Clostridium bacteria, Roseburia bacteria, Senegalimassilia anaerobia,* and others were found in much lower abundance in responders.

In the heatmap of microbial profiles, samples are clustered based on the similarity of their bacterial structures **(Fig. 3b)**. Interestingly, we found that the selected bacterial markers not only differentiate responders from non-responders but also distinguish between disease categories. For instance, PSC-UC samples are clustered together, as are PSC-CD samples, while non-responders, who were all PSC-alone, also form a distinct group. Samples from the same individuals are clustered together on both the heatmap **(Fig. 3b)** and the PCoA plot **(Fig. 3a)**, indicating a personalized gut microbiome, i.e., intra-individual differences are smaller than inter-individual ones. Therefore, disease categories can also be considered as a general predictor of LPD response.

### Gut metabolome is affected by multiple variables

To better understand metabolomic parameters of LPD response, we conducted the global untargeted metabolomic analysis of all stool samples using the ultra-performance liquid chromatography-tandem mass spectrometry (UPLC-MS) (see **Methods**). A total of 1,428 metabolites were quantified from at least one sample in this study, and a subset of 1,057 metabolites with known chemical names and corresponding identities in at least one of the following three databases: the Human Metabolome Database (HMDB) [33], the Kyoto Encyclopedia of Genes and Genomes (KEGG) database [34], and the PubChem database [35]. Metabolite compositions exhibited greater temporal variations within individuals than microbial profiles, as indicated by ordination analysis **(Fig. 3a** versus **Fig. 3c, Fig. S4a** versus **Fig. S4c)**, revealing that after dietary interventions, samples from the same individuals at different time points were interspersed with samples from others, leading to a mixed clustering pattern. Using the previously described multivariate linear regression model, we identified 29 metabolomic markers associated with clinical outcomes (p-value < 0.05 and q-value < 0.2) (see **Methods, Formula 1)**. Of these, 27 metabolites were positively associated with LPD responders, while 2 metabolites were negatively associated **(Fig. 3d)**. The intensity of metabolite markers in LPD responders versus LPD non-responders was less distinctive than that observed with microbial markers **(Fig. 3d)**. Additionally, in contrast to microbial markers, clustering based on metabolite markers composition did not show a clear correlation with disease category **(Fig. S6)**. Moreover, the observable intra-individual variation in these markers after dietary intervention indicates baseline metabolic profile cannot serve as a reliable predictor of response outcome to diet intervention, unlike microbial markers.

### Sulfur-related metabolites predict the LPD-induced changes

Diet-induced temporal changes in metabolite profiles were further analyzed to determine the primary mechanism underlying the effect of LPD. We employed multivariate linear regression models, which are different from those described earlier, to distinguish metabolite markers under LPD intervention (W4 and W8) from baseline (W0), incorporating the individual as a random effect and disease category, age, gender, and BMI as fixed effects **(see Methods, Formula 2)**. We identified 39 metabolite markers that significantly changed throughout LPD compared to baseline (p-value < 0.05 and q-value < 0.25), 21 of which have known chemical annotations. Of these, 6 (or 15) metabolites’ abundances increased (or decreased) with LPD. Notably, the majority of depleted metabolites (8/15) were either amino acids or fatty acids. Among the 6 enriched metabolites, four were classified as carbohydrates (n=1), medium-chain hydroxy acids (n=1), tricarboxylic acids (n=1), or quinoline carboxylic acids (n=1) (**Fig. 4a**).

**Figure 4.**
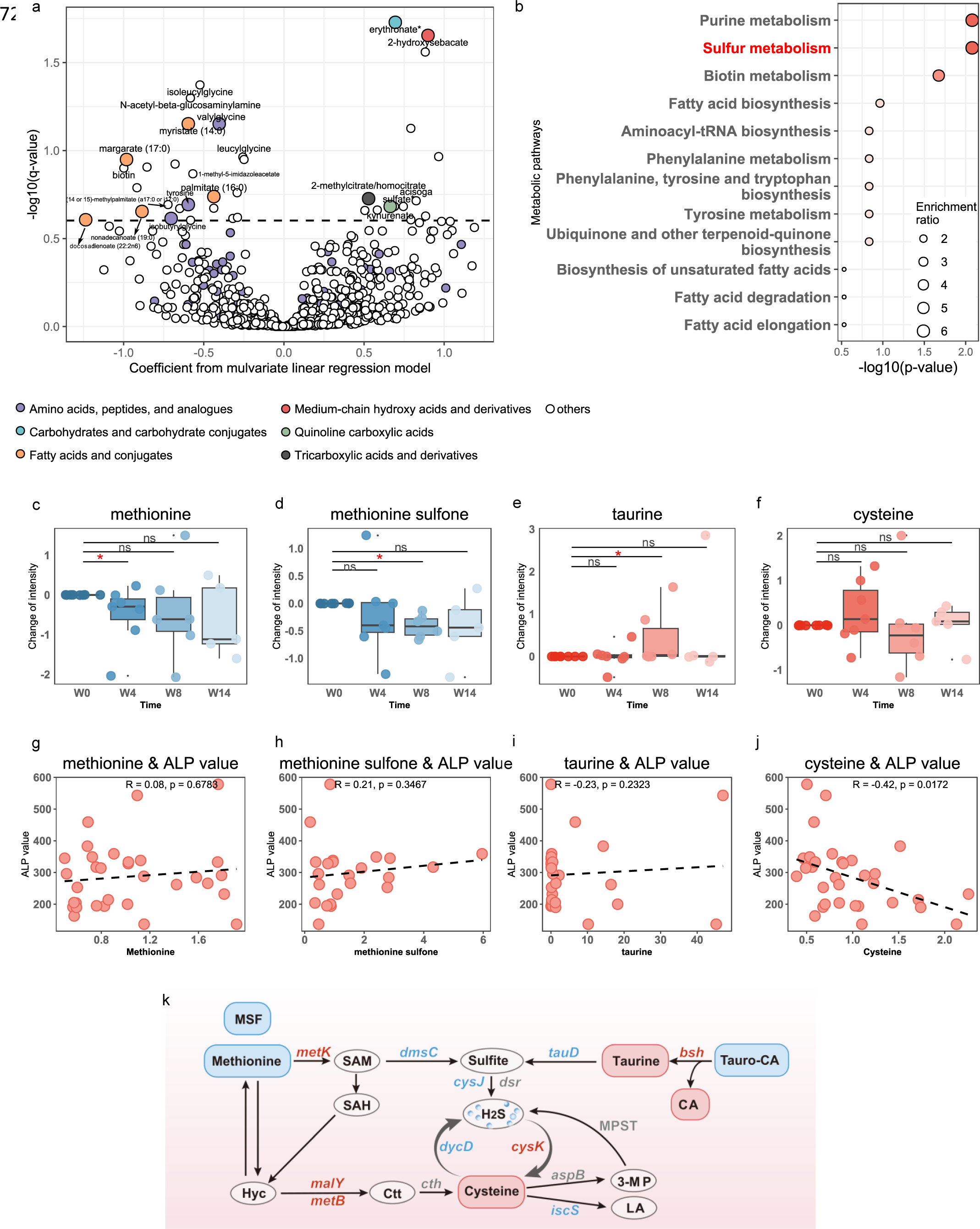
The fecal metabolomics predicts the disease improvement trajectory under LPD in those LPD responders. **a**) Volcano plot derived from multivariate linear regression models, using time series metabolite intensity profiles from W0, W4, and W8 and adjusting for covariates including disease categories. The metabolites were colored according to the class subclasses. **b**) Metabolite set enrichment analysis (MSEA) of selected metabolites with significant diet-induced changes after the regression model in Fig.3a (MetaboAnalyst v5.0, p-value < 0.05). **c-f**) Temporal changes in four key sulfur metabolism metabolites, including methionine, methionine sulfone, taurine, and cysteine. Bars are colored blue to represent metabolites that decreased after LPD, and orange for those that increased. The Wilcoxon signed-rank test was employed to assess significance between sampling time points, with * denoting significance (0.01< p-value < 0.05), and ‘ns’ indicating no significant difference (p-value > 0.05). **g-j**) The relationship between each of the four metabolites and ALP values tested using Spearman correlation. Cysteine shows a negative association with ALP values, whereas the other three metabolites have no significant association (p-value < 0.05). k) Characterized and proposed sulfur-related metabolic pathways in response to LPD intervention benefiting patients with PSC. Metabolites and genes that increased are colored red, those that decreased are blue, while those without significant changes are gray (Wilcoxon signed-rank test, p-value < 0.05). Abbreviations used are as follows: MSF: methionine-sulfone; Hyc: homocysteine; Ctt: cystathionine; LA: L-alanine; 3-MP: 3-Mercaptopyruvate; Tauro-CA: taurocholic acid; CA: cholic acid.

To identify metabolic pathways that are significantly altered, we performed metabolite set enrichment analysis (MSEA) [36, 37] using all the 21 metabolite features that significantly changed throughout treatment and with known chemical names. We found significant changes in purine metabolism (MetaboAnalyst v5.0, p-value = 0.0083), sulfur metabolism (p-value = 0.0083), and biotin metabolism (p-value = 0.021) with LPD. Notably, sulfur metabolism had the highest enrichment ratio (enrich = 6.26), suggesting it may be a primary mechanism behind the effect of LPD (**Fig. 4b**).

To extract metabolites potentially converted from or to sulfur-containing chemicals, out of the 1,043 detected metabolites, we identified 40 sulfur-related metabolites referenced in the SMILES database [38]. Four key metabolites were found to have significantly changed (**Fig. 4c-f & Fig. 4k**). Methionine (Wilcoxon signed-rank test, p-value=0.039 at W4) and its oxide methionine sulfone (Wilcoxon signed-rank test, p-value=0.015 at W8) were significantly depleted compared to baseline. This suggests that reduced protein intake led to a continual decrease in methionine and methionine sulfone in the gut. Taurine significantly enriched by W8 (Wilcoxon signed-rank test, p-value=0.047) compared to baseline. Cysteine level varied over time, increasing at W4 but decreasing at W8. Notably, cysteine intensity changes showed a significant negative association with ALP changes, indicating that cysteine accumulation correlates with lower ALP values (Spearman correlation: R = -0.42, p-value= 0.0172) (**Fig. 4j)**. Furthermore, in association with taurine accumulation, significant changes in luminal bile acids were observed. Notably, we found the levels of cholic acid was significantly elevated with LPD, while taurocholic acid were significantly decreased with LPD **(Fig. 4k & Fig. S7,** pathways referred to KEGG database [45]) A complete list of significantly changed sulfur-containing metabolites (n=45) and bile acids (n=19) is in **Table S1**.

### Changes in functional metagenomic features associated with LPD intervention

To explain the microbial contributions to the metabolite variations and to understand the temporal changes of functional consequences of the microbial community leading to response, we profiled gene families in all stool samples. Based on the shotgun metagenomic sequencing data, we analyzed the functional gene profiles using HUMAnN3.6 and identified 2,427 enzymes (KOs) from different bacterial species. Although the functional pathways were relatively stable across individuals and across times (**Fig. S8**), due to microbial functional redundancy, we then selected genes that significantly changed throughout dietary intervention in LPD responders by fitting a multivariate linear model to each enzyme, adjusting for the random effect of each individual and other covariates **(see Methods, Formula 2)**. In total, 99 (or 11)genes significantly increased (or decreased) at W4 and W8 compared to baseline (W0) (target p-value < 0.05).

We focused on genes involved with sulfur metabolism to decipher the relationship between sulfur-related metabolite changes and microbial functional changes. We referred to a comprehensive list of 74 genes involved in microbial sulfur metabolism in humans [39, 40]. Of these, 27 were detected in at least one sample in this study, with 6 (or 5) of these genes significantly decreasing (or increasing) during intervention in LPD responders (Wilcoxon rank-sum test, p-value < 0.05), and 4 genes increasing with marginal significance (0.05 < p-value < 0.06) (**Fig. 4k & Table S2**). Interestingly, LPD led to a reduced amount of *cysJ,* a critical enzyme for producing hydrogen sulfide (H_2_S) from sulfite, and a reduced amount of *dycD*, key for H_2_S production from cysteine **(Fig. 4k)** [39, 40]. Conversely, the *cysK* gene, involved in metabolizing H_2_S in the biosynthesis of cysteine, increased throughout LPD **(Fig. 4k)** [39, 40]. Similarly, there was an increase in the *bsh* gene that metabolizes taurine and aids in the deconjugation of tauro-conjugated bile acids excreted to the gut from the biliary tree **(Fig. 4k)** [41]. These findings highlight the relationship between microbial functional profile with metabolite changes, supporting the potential mechanistic role of altered sulfur metabolism in attenuating luminal inflammation and enhancing ALP reduction in PSC.

### Bacterial species associations with functional genes

Given the significant changes in microbial functional profiles, we explored the bacterial species connected to these enzymes. We constructed a large-scale association network among functional genes, metabolites, and bacterial species. To identify covariations strictly linked to diet change, we first residualized each feature in either measurement type using the same multivariate model employed to determine differential changes in responders during LPD intervention (see **Methods, Formula 2)**. This residualization process, using longitudinal measurements, minimizes inter-individual variation and highlights within-person associations over time, considering diagnostic categories as covariates. The resulting network contained 74 edges linking genes and metabolites, 906 edges between genes and species, and 57 edges linking gene pairs, where at least one connected gene was sulfur-related and the paired correlation was significant (Spearman correlation FDR < 0.05) (**Fig. 5a & Table S3**). The network encompassed 331 nodes spanning features from three measurement types. *tauD*, a gene converting taurine to sulfite (further convertible to H_2_S) **(Fig. 4k)**, was central in the network with the most connections (n = 174) to different bacterial species. However, for most of these species (90.2%), the abundance change of a single species was not significant (multivariate model p-value < 0.05). *Eggerthella lenta*, significantly positively associated with *tauD* (Spearman correlation FDR < 0.05) **(Table S3)**, showed a marked decrease in abundance during LPD intervention (Wilcoxon rank-sum test p-value = 8.7e-3; multivariate model: p-value = 0.016) **(Fig. 5d, Fig. S9 & Table S4)**. This species is critical in diet-induced changes, because it is also linked to *dmsC* (K00185), involved in the conversion from methionine to sulfite **(Fig. 4k)**, estimating via the taxonomic stratified functional profile by HUMAnNv3.6, and carries 10 other genes associated with sulfur metabolism, including *iscS*, and *ahcY*, which are genes significantly reduced with LPD (Wilcoxon rank-sum test, p-value < 0.05) as well **(Table S2)**. Genes *cysJ* and *dcyD* carried the second and fourth most connections, contributed by 92 and 74 different species, respectively (**Fig. 5a & Table S3**). These two genes were significantly interrelated (Spearman correlation coefficient = 0.95, R = 5.06e-11) and associated with other reduced genes including *tauD, dmsC, aspC,* and *iscC*. However, these genes were contributed by more than a single bacterium because no significant changes were found in any single species in *cysJ* or *dcyD* based on stratified abundance. *bsh*, a gene that metabolizes taurine and aids in the deconjugation of tauro-conjugated bile acids, did not exhibit positive correlations in any species in the complex association network. However, the taxonomic stratified analysis showed a significant increase in the abundance of *bsh* in *Roseburia intestinalis* in LPD responders (Wilcoxon rank-sum test, p-value = 1.7e-3), a decrease in the SCD group and no change in LPD non-responders **(Fig. 5b & Table S5)**. Notably, *Roseburia intestinalis* is a key bacterium in sulfur metabolism, containing 10 relevant genes that significantly changed, including *cysK, metK, dcm, metB, metY, luxS*, and others **(Fig. 5c & Table S5)**. Not only gene abundances, the bacterial abundance of *Roseburia intestinalis* showed a marked increase in LPD responders (Wilcoxon rank-sum test: p-value = 3.8e-3; multivariate model: p-value = 4.4e-4) **(Fig. 5d, Fig. S8 & Table S4)**.

**Figure 5.**
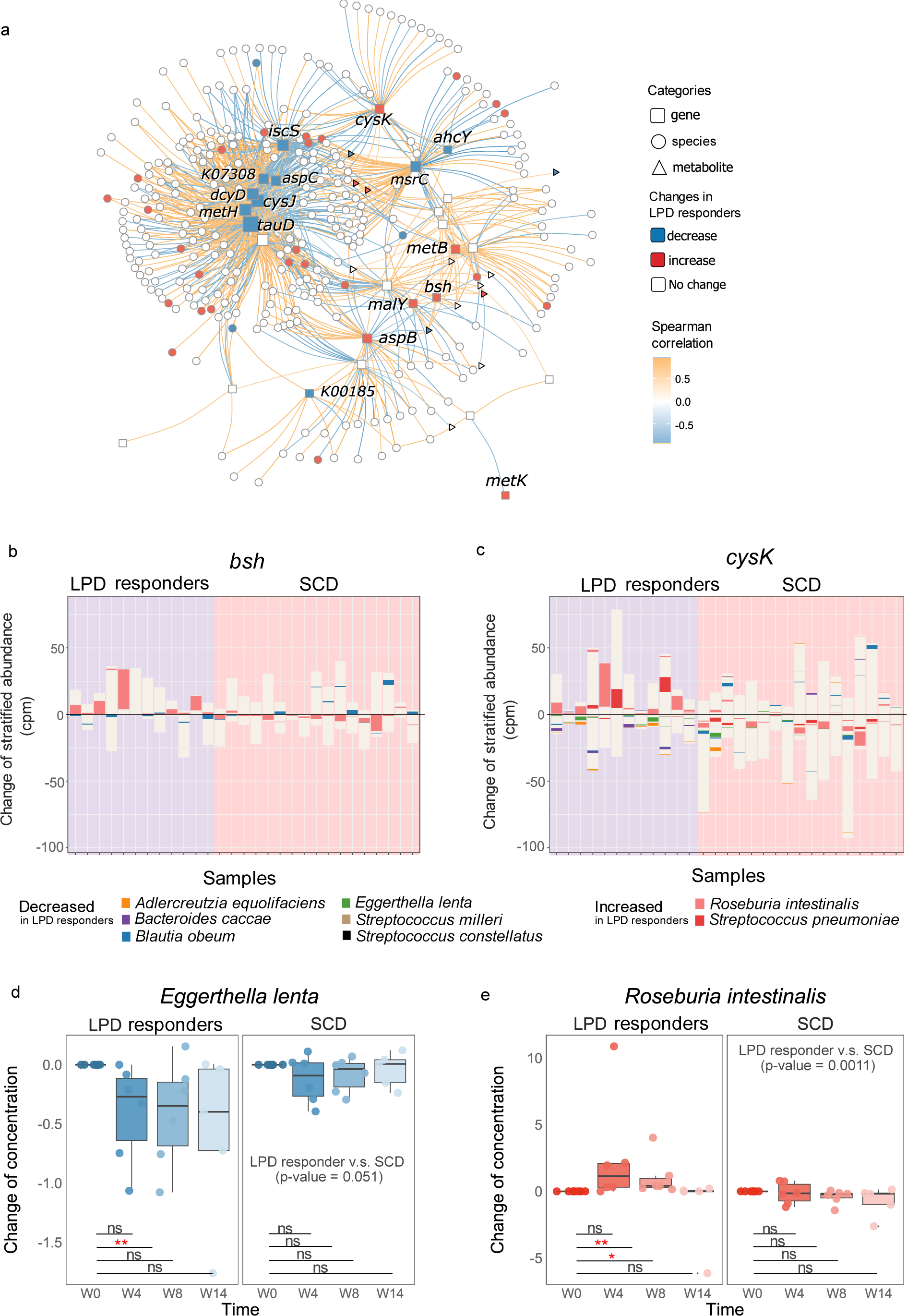
Microbial sulfur metabolism and associated bacterial species. **a**) Interactions among microbial sulfur metabolism genes, related metabolites, and relevant microbial species. Nodes were colored blue to represent features that decreased after LPD and red for those features that increased. Edge gradients represent the coefficients of Spearman correlation between the abundance (or intensity) of nodes. Only associations deemed significant, with FDR < 0.05, are shown here. **b-c**) Shifts in the stratified abundance of bacterial species for two represent genes (*bsh* and *cysK*) after dietary treatments. Each column is a sample, with the left panel on a purple background representing LPD responders under intervention, and the right panel on a pink background corresponding to samples from the SCD treatments. We subtracted the stratified abundance in each sample from its corresponding baseline to calculate the gene’s change amount. Each column contains stacked bars, with each bar representing the change in stratified abundance of a specific bacterial species identified as hosts of the specified genes. Values below the y=0 line indicate a decrease, while those above indicate an increase. Species are colored only if their stratified abundances showed significant changes (either increases or decreases) at W4 (or W8) compared to the baseline in LPD responders. The unit ‘cpm’ represents counts per million. **d-e**) Temporal changes of two key bacteria involved in sulfur metabolism: d) *Eggerthella lenta* and e) *Roseburia intestinalis*. Bars are colored blue to represent metabolites that decreased after LPD, and orange for those that increased. The Wilcoxon rank-sum test was employed to assess significance between sampling time points, with ** denoting very significant changes (0.001< p-value < 0.01), * significant (0.01< p-value < 0.05), and ‘ns’ indicating no significant difference (p-value > 0.05).

## DISCUSSION

The relationship between diet-microbiome-host interactions and their influence on the course of PSC is complex and poorly understood. To our knowledge, this is the first randomized controlled trial comparing the effects of two diets to assess a biochemical improvement in PSC through their modulatory effects on the gut microbiome. We observed that protein restriction was associated with improvement in ALP, while a diet limited in carbohydrates but not protein yielded no such benefit.

Moreover, the integration of multi-omics measurements, including state-of-the-art whole metagenomic sequencing and untargeted metabolite measurements, enabled us to reveal significant diet-induced changes in sulfur metabolism, purine metabolism, and biotin metabolism. Previous studies have suggested that H_2_S can be generated from multiple sulfur-containing amino acids, or from sulfate-reducing bacteria [19, 42]. In turn, H_2_S overproduction inhibits the cytochrome c oxidase activity in the human mitochondrial respiratory chain, thus disrupting the mitochondrial energy metabolism and promoting mucosal inflammation [43]. Although a potentially detrimental role of H_2_S has been linked to intestinal dysbiosis, much less is known about its relationship to liver disease progression. Our study provides evidence of significant diet-induced changes in sulfur-containing metabolites and related enzymes. Notably, decreased protein intake led to reduced levels of methionine and its oxide, associated with lower microbial enzymatic activity, particularly in enzymes involved in H_2_S production. We also observed a decrease in H_2_S-producing bacteria, including *Eggerthella lenta*, significantly associated with enzymes like *tauD, dmsC, iscS*, and *ahcY*. Thus, our data confirm, from various perspectives, that a low-protein diet regulates H_2_S production by correcting the imbalance in sulfur metabolism.

Alterations in sulfur metabolism as the primary mechanism for the beneficial effects of low protein are also reflected in the accumulation of taurine and cysteine. This accumulation results from increased activity of the *bsh* gene, associated with taurine accumulation, and the *malY* and *metB* genes, linked to cysteine production, when the conversion of taurine and cysteine to H_2_S was significantly reduced, as discussed previously. An unhealthy gut microbiome that lacks the required deconjugation of host primary bile acids into secondary bile acids, impairs liver and gut function, in addition to overall health [44, 45]. Importantly, our data showed that taurine accumulation and cholate increase correspond with the increase of deconjugation genes, such as *bsh*, in response to taurocholate—a taurine-conjugated bile acid, indicating amelioration of the imbalanced gut microbiome in PSC patients with restricted protein intake. Intriguingly, the bacteria found to increase after LPD, primarily carriers of *bsh, cysK, malY*, and *metB* genes, include *Roseburia intestinalis*. The increase in *Roseburia intestinalis,* known for its beneficial role in producing beneficial short-chain fatty acids which possess anti-inflammatory properties [46, 47], suggests a rebalancing of the bacterial structure in PSC patients, where deconjugation is otherwise lacking compared with healthy individuals. Therefore, our data suggest that LPD might enhance the gut’s deconjugation of primary bile acids, offering potential protective mechanisms against inflammation and helping to restore a healthier microbial balance/composition.

While the role of H_2_S has not been previously investigated in PSC, more has been done to investigate its role in UC with a small but largely supportive literature [48, 49]. An increase in sulfate-reducing bacteria has been associated with UC and flares in particular [50, 51]. With regard to diet, flares have been found to be associated with a higher protein diet [52]. Higher protein diets have been demonstrated to increase H_2_S production by intestinal bacteria increasing sulfate-reducing bacteria [53]. Several interventional studies of a low sulfur diet have shown to be of benefit in UC though more detailed investigations into the metagenomic and metabolomic elements and pathologic pathways have not been part of those studies [54]. Still, the intimate connection between UC and PSC would suggest these studies might be relevant and H_2_S may offer a potential physiologic link between these diseases.

In addition to our findings regarding sulfur metabolism, we were able to identify microbial signatures that can be used to predict response to LPD. Besides, we observed that the majority of responders had concomitant UC, and that the predictive microbial signatures were enriched in all baseline samples of PSC-UC participants. Collectively, these findings serve to enhance patient selection for this intervention and warrant further exploration in larger cohorts.

Although this study provides significant insights into better understanding PSC pathophysiology, there are several limitations, including a small cohort size and lack of a traditional control arm. Future inclusion of healthy controls or IBD-only controls could offer a more comprehensive understanding of mechanisms involved in disease progression. Additionally, while the 8-week-long intervention and 4-week self-guided follow-up provided interesting findings, exploring the longer-term effects and potential changes after prolonged intervention remains a key area for future research.

Overall, the well-documented dietary adherence and records of different subgroups of individuals with PSC, in combination with high-throughput technologies and the multi-omics approach, enabled us to determine the clinical, microbial and metabolic signatures associated with responsiveness to dietary interventions. This study enhanced our understanding of PSC pathogenesis and paved the way for large-scale dietary intervention studies in PSC.

## METHODS

### Study setting and design

This was a decentralized randomized controlled trial across the United States. Participant enrollment occurred between August 28, 2020 and June 9, 2021. Recruitment occurred in-person and by video remotely, and participants were able to conduct the study remotely. Recruitment was facilitated via physician referral, patient advocacy organizations (PSC Partners Seeking a Cure and Consortium for Autoimmune Liver Disease), and self-referral.

Eligible participants were randomized in 1:1 fashion to the LPD and SCD at a screening visit (week 0, W0, referred to as “baseline”) during which medical records were reviewed, eligibility was assessed and bloodwork, stool, baseline Food Frequency Questionnaire (FFQ) and Patient-reported Outcome Measures (PROMs) were collected. Thereafter, participants were given two weeks for education on appropriate guidelines for the assigned dietary intervention as well as procurement of food. The dietary intervention began at week 2 (W2) and lasted for 8 consecutive weeks until week 10 (W10) under dietitian supervision, with an option to continue self-directed for an additional 4 weeks (W14). There was a total of 7 video visits with a research dietitian. Additional dietary counselling was also offered between screening and baseline visits to provide further instruction and support. Three-day food diaries, bloodwork, stool samples and PROMs were collected at 7 points throughout the study (**Fig. 1a**).

### Eligibility criteria

Adults between 18 and 70 years of age with large-duct PSC diagnosed by typical cholangiogram findings with no evidence of a secondary cause of sclerosing cholangitis and with serum ALP >1.5 times the upper limit of normal (ULN) were potentially eligible for enrollment. Participants with concomitant UC or CD were eligible if the Simple Clinical Colitis Activity Index or Harvey-Bradshaw Index, respectively, were <5. Additional inclusion criteria included platelet count ≥ 150,000/mm^3^, serum albumin ≥ 3.3 g/dL, serum creatinine < ULN, and stable dose or no use of ursodeoxycholic acid (UDCA) for at least 3 months prior to enrollment for those taking and not taking UDCA respectively. Proficiency in English and ability to complete PROMs independently was also a requirement. Exclusion criteria included pregnancy or lactation, ALT above 10 times the ULN, total bilirubin at least twice the ULN, INR >1.2, decompensated cirrhosis, small duct PSC, other etiologies of liver disease, positive AMA, history of liver transplantation, history of hepatocellular carcinoma or cholangiocarcinoma, ascending cholangitis within 90 days of enrollment, antibiotic use within 6 weeks prior to enrollment or planned during the study period, current vegetarian or adherence to the SCD, nut allergy given that nut flour is a dietary staple of many SCD recipes and nut allergy could compromise diet adherence, celiac disease, history of malignancy within 5 years with the exception of adequately treated cervical carcinoma in situ and basal or squamous cell carcinoma, inability to complete a dietary log, or concurrent participation in another therapeutic clinical trial. Medical records of all individuals were reviewed at the screening visit to determine eligibility.

### Dietary interventions

The LPD was developed according to 2015-2020 USDA Dietary Guidelines of a vegan diet which state the diet should be rich in grains, legumes, nuts and other plant-based proteins, with a focus on increased consumption of fruits, vegetables and healthy fats [55]. In addition to these guidelines, the diet restricted high-sulfate items, defined as containing >100mg sulfur per 100g of food item. A typical vegan diet contains approximately 2.3g/day of sulfur-containing amino acids, falling within the estimated range of appropriate sulfur content of 2.1-3.0 g/day [56]. The SCD followed the dietary guidelines detailed in the book *Breaking the Vicious Cycle* by Elaine Gottschall [57]. The detailed LPD and SCD dietary guidance is provided in **Table S6**. Participants were provided with education materials, recipes and a food procurement stipend for their assigned diet at the screening visit. Total daily energy intake was calculated by the Mifflin St. Jeor equation using an activity factor of 1.3 reflecting light activity and exercise level.

### Assessment of diet composition and adherence

Habitual diet was recorded using a validated FFQ at the screening visit (week 0) that queried dietary habits over the preceding year. During the intervention phase, 3-day food diaries were recorded at five time points (weeks 2, 4, 6, 8, 10) (**Fig. 1a**). One of these days included a weekend. Participants recorded diet in real-time via photodocumentation of each meal, snack and beverage using a smartphone application that was customized for this study. The application transmitted time-stamped photographs taken during the recording period, along with a reference-sized study ruler to assess relative sizes of food items. Caloric intake and micronutrient composition of each meal was calculated and analyzed using Food Processor, a program with the ability to quantify protein intake, as well as cysteine, methionine and taurine content as a proxy for sulfur.

In order to optimize compliance, the research team reviewed dietary records in real-time and discussed barriers to compliance with participants when appropriate. Dietary analysis was also performed at the conclusion of each diet recording period to further assess compliance. Feedback was provided to participants via the application if there were fewer than 2 entries per day or if a 24-hour period elapsed without any recording. The study coordinator followed up by email and/or telephone if no confirmation was received from the participant within 24 hours of the communication.

### Outcome measures

The outcome measures were response in alkaline phosphatase (ALP), Alanine Aminotransferase (ALT) and Aspartate Aminotransferase (AST).

### Whole Metagenomic Sequencing

Stool samples were collected at home frozen within 15 minutes of collection and maintained at −80 ℃ before sending to Diversigen for DNA extraction, whole-genome shotgun library preparation, and Illumina sequencing. The metagenomics sequencing targeted approximately 5 Gb of sequences per sample, utilizing 151 base pair paired-end reads.

### Read-level quality control and metagenomic profiling

Raw sequencing reads underwent quality control (QC) using KneadData version 0.12.0 (https://huttenhower.sph.harvard.edu/kneaddata/). Briefly, the first step of this process utilizes Trimmomatic version 0.39 for forward/reverse adapter removal, low-quality reads trimming and tandem repeats removal using the default parameters. Contaminants from the host (human) genomes were subsequently identified and removed by mapping against the reference database (hg37_and_human_contamination) using Bowtie2 version 2.5.1.

After QC, each dataset contained over 2.8 Gb of clean, paired-end reads. Clean reads were then taxonomically profiled using MetaPhlAn4 version 4.0.6 [58], which includes an updated database significantly larger than the previous version 3.1. Species-level relative abundances were considered all this study. Species that failed to exceed 0.1% average relative abundance or were detected in less than 4 samples were excluded. Functional profiling was performed using HUMAnN3 version 3.6 under default parameters according to the UniRef90 definition to get the relative abundance of gene families in the unit of reads per kilobase (RPK) [59]. And MetaCyc provides pathway definitions to group gene families to get pathway abundance and coverage. The abundances of gene families were further transformed into the unit of copies per million (CPM), and we regrouped the gene families to KEGG Orthogroups (KOs), to filter out 74 gene families that were involved in the microbial sulfur transformation.

### Global untargeted metabolomics

Stool samples were sent to Metabolon for global untargeted metabolomics following the standard procedure. Briefly, samples were pretreated to remove proteins and recover chemically diverse metabolites. The resulting extract was divided into five fractions: two for analysis by two separate reverse phases (RP)/UPLC-MS/MS methods with positive ion mode electrospray ionization (ESI), one for analysis by RP/UPLC-MS/MS with negative ion mode ESI, one for analysis by HILIC/UPLC-MS/MS with negative ion mode ESI, and one sample was reserved for backup. All methods utilized a Waters ACQUITY ultra-performance liquid chromatography (UPLC) and a Thermo Scientific Q-Exactive high resolution/accurate mass spectrometer interfaced with a heated electrospray ionization (HESI-II) source and Orbitrap mass analyzer operated at 35,000 mass resolution. Raw data were extracted, peak-identified and QC processed using the contractor’s hardware and software. Compounds were identified by comparison to library entries of purified standards or recurrent unknown entities. The intensity of each compound was quantified using area-under-the-curve.

### Metabolite-level quality control and pretreatment

Metabolite intensities were normalized as z-scores: 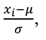, where, *X^i^* represents the original metabolite intensity, *μ* is the mean value of that metabolite intensity across all samples, and *σ* is the standard deviation of that metabolite intensity across all samples. Subsequently, missing values were imputed using the minimum normalized value for each compound. The normalized and imputed values were used in the association and multivariate linear regression analyses throughout the context. With the exception of the estimation of temporal changes of metabolite intensity in each individual, the original intensities of specified metabolites in W0 were subtracted from their original intensity in W4 (or W8, W14).

### Statistical analyses

Prior to downstream analysis, metagenomic and metabolomic were combined. Samples with complete profiles of both types were used in the downstream analysis, totalling 64 samples. Three samples with only metabolomics datasets were excluded. From a total of 108 nutrients, 59 nutrient features were selected based on domain knowledge. These features, measured in various units, were log-transformed before we adjusted them for total energy intake to get the nutrient densities. This was followed by Lasso regression utilizing the ‘glmnet’ library in RStudio to select significant dietary features (predictors) that are non-zero [60]. The response variable was the corresponding ALP value. Subsequently, we incorporated the selected features and included patient ages as an additional variable to construct a linear model. The coefficients and p-values were extracted for statistical significance. For each patient, the Change of ALP value is estimated by subtracting the baseline ALP (W0) from the ALP at W4 (or W8, W14), then dividing by the baseline ALP (W0). Then for each disease category (PSC-UC/PSC-CD/PSC-alone), the Potential to Response was further calculated as the number of samples with reduced ALP value divided by the total number of samples. Alpha diversity of gut microbiota was estimated using the Shannon index and microbial richness (detected species counts). Principal coordinates analysis was performed using the bray-curtis distance of the relative abundance of species in all samples. Metabolite set enrichment analysis (MSEA) was conducted quantitatively through the online platform of MetaboAnalyst 5.0 [36].

### Linear regression models

We used multivariate linear regression models to identify potential microbial markers distinguishing responders from non-responders after LPD. Abundances were fitted with the following species-specific linear mixed-effects model,

> feature ∼ **response outcomes** + diagnostic categories + gender + age +BMI + (1 | individual) + ε
>
> **(Formula 1)**

In each species-specific multivariate model, the abundance of each species was modeled as the function of the binary response outcomes (responders/non-responders, with non-responder as reference) within each individual (as random effect), while adjusting for diagnostic categories (PSC-UC/PSC-CD/PSC-alone, with PSC-alone as reference), genders (male/female, with male as reference), age (continuous variable) and BMI (continuous variable). MaAsLin2 [61] in RStudio was the package used to fit the model, with p-values adjusted for multiple hypothesis testing and a target FDR of 0.2.

Separately, another series of multivariate linear regression models were fitted to find a differential abundance of features after LPD versus baseline using microbial profile, metabolic profile, and microbial functional profile, respectively. The model is as below,

> feature ∼ **treatment stages** + diagnostic categories + gender + age +BMI + (1 | individual) + ε
>
> **(Formula 2)**

The difference between this model and the previous one is that the abundance of each feature is modeled as the function of the treatment stage (baseline/after LPD, with baseline as reference) instead of response results, while the rest fixed effect and random effect are not changed. The baseline corresponds to samples in W0, while after LPD corresponds to samples in W4 and W8.

Only those with a response of ALP were considered to fit these models to explain the dietary effects of the LPD.

## Clinicaltrials.gov ID

NCT04678219

## Author Contributions

JK,YYL, GS, DB, MA, LG, FS, ES, and CB designed the project. GS led the clinical trial. XY led the data analysis and manuscript preparation. JK, YYL, GS, ZS, SK, DB, SDM, MA, SH, TC, MP, NJ, MD, AA, LG, FS, ES, and CB interpreted the results. XY prepared the manuscript. GS, JK, and YYL edited the manuscript. All authors reviewed and approved the manuscript. JK and YYL supervised the study.

## Data Availability

All data produced in the present study are available upon reasonable request to the authors

## Acknowledgements.

This work was supported by the Resnek Family Center for PSC Research.

## REFERENCES

1. Hov, J.R. and T.H. Karlsen, The microbiota and the gut-liver axis in primary sclerosing cholangitis. Nat Rev Gastroenterol Hepatol, 2022.

2. Sayed, A., et al., Predictors and Outcomes of Biologic/Immunosuppressive Therapy for PSC-Associated IBD. Gastroenterology, 2022. 162(3): p. S76.

3. Mertz, A., et al., Primary sclerosing cholangitis and inflammatory bowel disease comorbidity: an update of the evidence. Ann Gastroenterol, 2019. 32(2): p. 124–133.

4. Hsu, C.L. and B. Schnabl, The gut-liver axis and gut microbiota in health and liver disease. Nat Rev Microbiol, 2023. 21(11): p. 719–733.

5. Jiang, X. and T.H. Karlsen, Genetics of primary sclerosing cholangitis and pathophysiological implications. Nature Reviews Gastroenterology & Hepatology, 2017. 14(5): p. 279–295.

6. Hov, J.R. and T.H. Karlsen, The microbiota and the gut–liver axis in primary sclerosing cholangitis. Nature Reviews Gastroenterology & Hepatology, 2023. 20(3): p. 135–154.

7. Timur, L., et al., Alterations of the bile microbiome in primary sclerosing cholangitis. Gut, 2020. 69(4): p. 665.

8. Vieira-Silva, S., et al., Quantitative microbiome profiling disentangles inflammation-and bile duct obstruction-associated microbiota alterations across PSC/IBD diagnoses. Nat Microbiol, 2019. 4(11): p. 1826–1831.

9. Fitzpatrick, J.A., et al., Dietary management of adults with IBD - the emerging role of dietary therapy. Nat Rev Gastroenterol Hepatol, 2022. 19(10): p. 652–669.

10. Lloyd-Price, J., et al., Multi-omics of the gut microbial ecosystem in inflammatory bowel diseases. Nature, 2019. 569(7758): p. 655-662.

11. Federici, S., et al., Targeted suppression of human IBD-associated gut microbiota commensals by phage consortia for treatment of intestinal inflammation. Cell, 2022. 185(16): p. 2879–2898 e24.

12. Zhang, Y., et al., Discovery of bioactive microbial gene products in inflammatory bowel disease. Nature, 2022. 606(7915): p. 754-760.

13. Hov, J.R. and M.J.C.O.i.G. Kummen, Intestinal microbiota in primary sclerosing cholangitis. 2016. 33: p. 85–92.

14. Kentaro, I., et al., Characterisation of the faecal microbiota in Japanese patients with paediatric-onset primary sclerosing cholangitis. Gut, 2017. 66(7): p. 1344.

15. Nakamoto, N., et al., Gut pathobionts underlie intestinal barrier dysfunction and liver T helper 17 cell immune response in primary sclerosing cholangitis. Nature Microbiology, 2019. 4(3): p. 492–503.

16. Kummen, M., et al., Altered Gut Microbial Metabolism of Essential Nutrients in Primary Sclerosing Cholangitis. Gastroenterology, 2021. 160(5): p. 1784–1798 e0.

17. Liu, Q., et al., Altered faecal microbiome and metabolome in IgG4-related sclerosing cholangitis and primary sclerosing cholangitis. Gut, 2022. 71(5): p. 899–909.

18. Ichikawa, M., et al., Bacteriophage therapy against pathological Klebsiella pneumoniae ameliorates the course of primary sclerosing cholangitis. Nat Commun, 2023. 14(1): p. 3261.

19. Wolfson, S.J., et al., Bacterial hydrogen sulfide drives cryptic redox chemistry in gut microbial communities. Nature Metabolism, 2022. 4(10): p. 1260–1270.

20. Barton, L.L., et al., Sulfur Cycling and the Intestinal Microbiome. Digestive Diseases and Sciences, 2017. 62(9): p. 2241–2257.

21. Pitcher, M.C.L., E.R. Beatty, and J.H. Cummings, The contribution of sulphate reducing bacteria and 5-aminosalicylic acid to faecal sulphide in patients with ulcerative colitis. Gut, 2000. 46(1): p. 64.

22. Próchnicki, T., et al., Mitochondrial damage activates the NLRP10 inflammasome. Nature Immunology, 2023. 24(4): p. 595–603.

23. Lewis, J.D., et al., A Randomized Trial Comparing the Specific Carbohydrate Diet to a Mediterranean Diet in Adults With Crohn’s Disease. Gastroenterology, 2021. 161(3): p. 837–852.e9.

24. Cohen, S.A., et al., Clinical and mucosal improvement with specific carbohydrate diet in pediatric Crohn disease. J Pediatr Gastroenterol Nutr, 2014. 59(4): p. 516–21.

25. Wastyk, H.C., et al., Gut-microbiota-targeted diets modulate human immune status. Cell, 2021. 184(16): p. 4137–4153.e14.

26. Slomski, A., Mediterranean Diet vs Low-fat Diet for Patients With Heart Disease. Jama, 2022. 327(24): p. 2386.

27. Longo, V.D. and R.M. Anderson, Nutrition, longevity and disease: From molecular mechanisms to interventions. Cell, 2022. 185(9): p. 1455–1470.

28. Kahleova, H., S. Levin, and N.D. Barnard, Vegetarian Dietary Patterns and Cardiovascular Disease. Prog Cardiovasc Dis, 2018. 61(1): p. 54–61.

29. Gentile, C.L. and T.L. Weir, The gut microbiota at the intersection of diet and human health. Science, 2018. 362(6416): p. 776-780.

30. Corbin, K.D., et al., Host-diet-gut microbiome interactions influence human energy balance: a randomized clinical trial. Nat Commun, 2023. 14(1): p. 3161.

31. Bajaj, J.S., S.C. Ng, and B. Schnabl, Promises of microbiome-based therapies. Journal of Hepatology, 2022. 76(6): p. 1379–1391.

32. Liwinski, T., et al., A prospective pilot study of a gluten-free diet for primary sclerosing cholangitis and associated colitis. Aliment Pharmacol Ther, 2023. 57(2): p. 224–236.

33. Wishart, D.S., et al., HMDB 5.0: the Human Metabolome Database for 2022. Nucleic Acids Res, 2022. 50(D1): p. D622–d631.

34. Kanehisa, M., et al., KEGG for taxonomy-based analysis of pathways and genomes. Nucleic Acids Res, 2023. 51(D1): p. D587–d592.

35. Kim, S., et al., PubChem 2023 update. Nucleic Acids Research, 2022. 51(D1): p. D1373-D1380.

36. Pang, Z., et al., Using MetaboAnalyst 5.0 for LC–HRMS spectra processing, multi-omics integration and covariate adjustment of global metabolomics data. Nature Protocols, 2022. 17(8): p. 1735–1761.

37. Xia, J. and D.S. Wishart, MSEA: a web-based tool to identify biologically meaningful patterns in quantitative metabolomic data. Nucleic Acids Research, 2010. 38(suppl_2): p. W71-W77.

38. Weininger, D., et al., SMILES. 2. Algorithm for generation of unique SMILES notation. 1989. 29(2): p. 97–101.

39. Wolf, P.G., et al., Diversity and distribution of sulfur metabolic genes in the human gut microbiome and their association with colorectal cancer. Microbiome, 2022. 10(1): p. 64.

40. Marcelino, V.R., et al., Disease-specific loss of microbial cross-feeding interactions in the human gut. Nat Commun, 2023. 14(1): p. 6546.

41. Heinken, A., et al., Systematic assessment of secondary bile acid metabolism in gut microbes reveals distinct metabolic capabilities in inflammatory bowel disease. Microbiome, 2019. 7(1): p. 75.

42. Lobel, L., et al., Diet posttranslationally modifies the mouse gut microbial proteome to modulate renal function. 2020. 369(6510): p. 1518-1524.

43. Blachier, F., M. Beaumont, and E. Kim, Cysteine-derived hydrogen sulfide and gut health: a matter of endogenous or bacterial origin. Curr Opin Clin Nutr Metab Care, 2019. 22(1): p. 68–75.

44. Guzior, D.V. and R.A. Quinn, Review: microbial transformations of human bile acids. Microbiome, 2021. 9(1): p. 140.

45. Collins, S.L., et al., Bile acids and the gut microbiota: metabolic interactions and impacts on disease. Nature Reviews Microbiology, 2023. 21(4): p. 236–247.

46. Xing, K., et al., <em>Roseburia intestinalis</em= generated butyrate boosts anti-PD-1 efficacy in colorectal cancer by activating cytotoxic CD8<sup=+</sup= T cells. Gut, 2023. 72(11): p. 2112.

47. Nie, K., et al., Roseburia intestinalis: A Beneficial Gut Organism From the Discoveries in Genus and Species. 2021. 11.

48. Stummer, N., et al. Role of Hydrogen Sulfide in Inflammatory Bowel Disease. Antioxidants, 2023. 12, DOI: 10.3390/antiox12081570.

49. Teigen, L.M., et al. Dietary Factors in Sulfur Metabolism and Pathogenesis of Ulcerative Colitis. Nutrients, 2019. 11, DOI: 10.3390/nu11040931.

50. Rowan, F.E., et al., Sulphate-reducing bacteria and hydrogen sulphide in the aetiology of ulcerative colitis. British Journal of Surgery, 2009. 96(2): p. 151–158.

51. Khalil, N.A., et al., In vitro batch cultures of gut microbiota from healthy and ulcerative colitis (UC) subjects suggest that sulphate-reducing bacteria levels are raised in UC and by a protein-rich diet. International Journal of Food Sciences and Nutrition, 2014. 65(1): p. 79–88.

52. Jowett, S.L., et al., Influence of dietary factors on the clinical course of ulcerative colitis: a prospective cohort study. Gut, 2004. 53(10): p. 1479.

53. Magee, E.A., et al., Contribution of dietary protein to sulfide production in the large intestine: an in vitro and a controlled feeding study in humans123. The American Journal of Clinical Nutrition, 2000. 72(6): p. 1488–1494.

54. Day, A.S., et al., Therapeutic Potential of the 4 Strategies to SUlfide-REduction (4-SURE) Diet in Adults with Mild to Moderately Active Ulcerative Colitis: An Open-Label Feasibility Study. The Journal of Nutrition, 2022. 152(7): p. 1690–1701.

55. U.S. Department of Health and Human Services and U.S. Department of Agriculture. 2015 – 2020 Dietary Guidelines for Americans. 8th Edition. December 2015.

56. Tuttle, S.G., et al., Further Observations on the Amino Acid Requirements of Older Men: II. Methionine and Lysine. The American Journal of Clinical Nutrition, 1965. 16(2): p. 229–231.

57. Gottschall, E., Breaking the vicious cycle: intestinal health through diet. 1994: Kirkton, Ont.: Kirkton Press.

58. Blanco-Míguez, A., et al., Extending and improving metagenomic taxonomic profiling with uncharacterized species using MetaPhlAn 4. Nature Biotechnology, 2023. 41(11): p. 1633–1644.

59. Beghini, F., et al., Integrating taxonomic, functional, and strain-level profiling of diverse microbial communities with bioBakery 3. eLife, 2021. 10: p. e65088.

60. Friedman, J.H., T. Hastie, and R. Tibshirani, Regularization Paths for Generalized Linear Models via Coordinate Descent. Journal of Statistical Software, 2010. 33(1): p. 1–22.

61. Mallick, H., et al., Multivariable association discovery in population-scale meta-omics studies. PLOS Computational Biology, 2021. 17(11): p. e1009442.

