## Supplementary material for "Demonstrating the Beneficial Effect of Low Protein Diet in Primary Sclerosing Cholangitis through a Randomized Clinical Trial and Multi-omics Data Analysis": SI.pdf

### Supplementary figures

**Fig. S1.** Percent change in Aspartate Aminotransferase (AST) and Alanine Aminotransferase (ALT) from the baseline before, during and after dietary intervention. Each line represents an individual.

**Fig. S2.** Carbohydrate intake of each individual during the 14 weeks of observation.

**Fig. S3.** Diagnostic categories (PSC-UC, PSC-CD, and PSC-alone) showed diverse microbiota, revealed by a) microbial richness (the number of detected species), b) Shannon index, and c) beta diversity.

**Fig. S4.** Temporal variations of microbial alpha diversity revealed by the richness and Shannon index in each arm.

**Fig. S5**. Beta-diversity of bacterial species (a and b) and metabolites (c and d) across both arms (LPD and SCD).

**Fig. S6.** Heatmap of metabolite markers that can distinguish LPD responders from non-responders, as determined by multivariate linear regression models (target p-value < 0.05 and FDR < 0.2).

**Fig. S7.** Temporal changes in key bile acids, including cholate, and taurocholate (also named taurocholic acid). Bars are colored blue to represent metabolites that decreased after LPD, and orange for those that increased.

**Fig. S8.** Functional pathway classes across individuals and across time obtained from HUMAnN3.6 and referenced to the MetaCyc database.

**Fig. S9.** Bacterial species exhibiting significant changes compared to baseline levels at Week 4 and Week 8 after LPD treatment, as revealed by a multivariate linear regression model (Formula 2, targeting p-values < 0.05).

### Supplementary tables

**Table S1**. A list of significantly changed sulfur-containing metabolites (n=18) and bile acids (n=9), comparing each metabolite intensity at W4 or W8 to its baseline level (only LPD responders were included) revealed by the Wilcoxon signed-rank test. (In a separate file)

**Table S2.** Genes in microbial sulfur metabolism detected in this study and their changes from baseline level at W4 or W8 (only LPD responders were included), or differences in LPD responders from the SCD arm. (In a separate file)

**Table S3.** Association network among functional genes, metabolites, and bacterial species (Spearman correlation targeting FDR < 0.05). (In a separate file)

**Table S4.** Bacterial species with significant changes compared to baseline level at Week 4 and Week 8, revealed by a multivariate linear regression model (Formula 2, targeting p-value < 0.05). (In a separate file)

**Table S5.** Taxonomic stratified functional profiles reveal the abundance of specific genes carried by a specific bacterial species. Here are genes with significant changes from baseline level at W4 and W8 (only LPD responders were included). (In a separate file)

**Table S6.** Detailed guidance for the LPD and SCD diets.

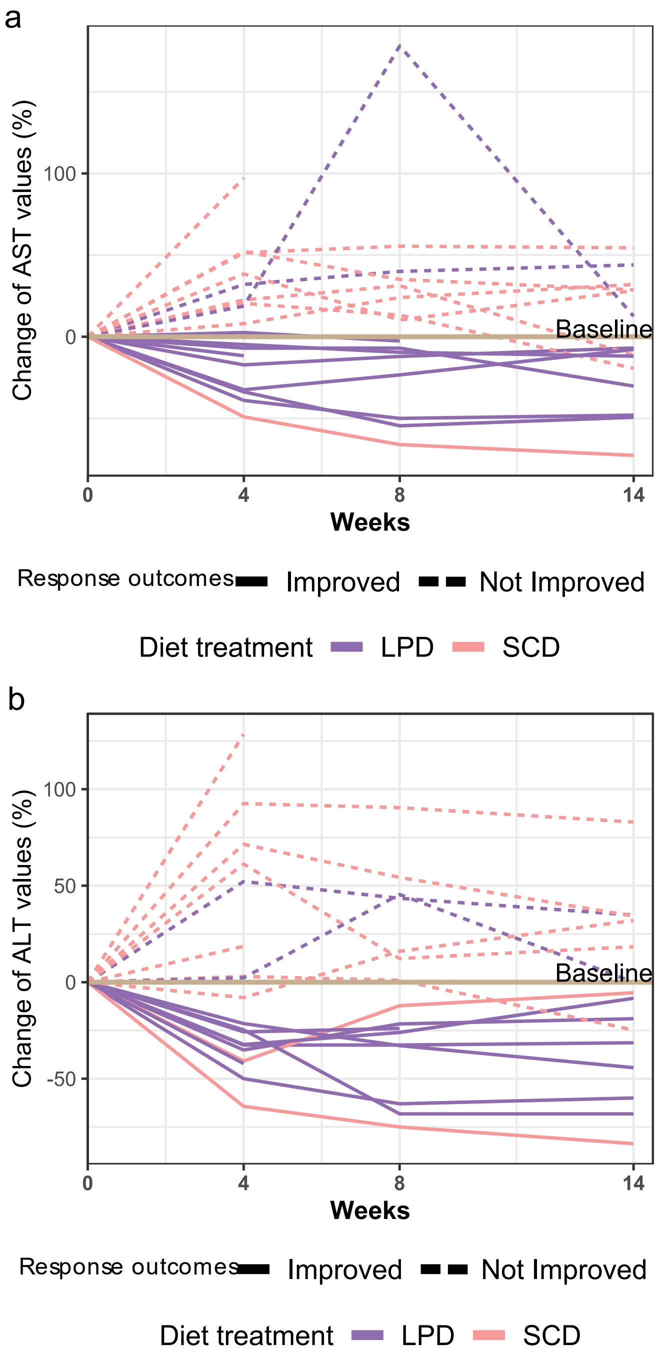


**Fig. S1.** Percent change in Aspartate Aminotransferase (AST) and Alanine Aminotransferase (ALT) from the baseline before, during and after dietary intervention. Each line represents an individual.

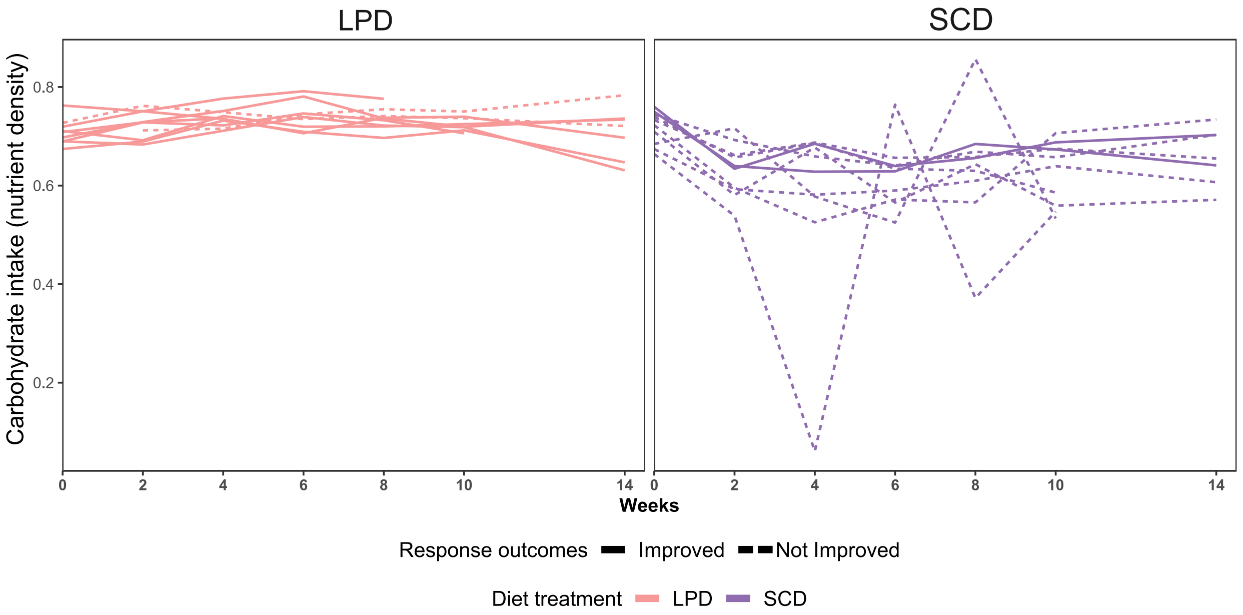


**Fig. S2.** Carbohydrate intake of each individual during the 14 weeks of observation. There were no significant differences between LPD and SCD at week 0 (Mann-Whitney U test, p-value = 0.549). After the dietary intervention spanning from Week 2 to Week 10, the levels in the SCD group were significantly lower than those in the LPD group at the corresponding time point (Wilcoxon rank-sum test, p-values are 0.00065, 2.17e-5, 0.0014, 0.0031 and 0.00050 for week 2, week 4, week 6, week 8 and week 10, respectively). Regarding differences within the SCD group, carbohydrate intake in SCD changed sharply from corresponding baseline levels after intervention at Week 2 till Week 10. The corresponding p-values between Weeks 2, 4, 6, 8, 10 and Week 0 are 0.0039, 0.0019, 0.0098, 0.064 and 0.0020, respectively (Wilcoxon signed-rank test).

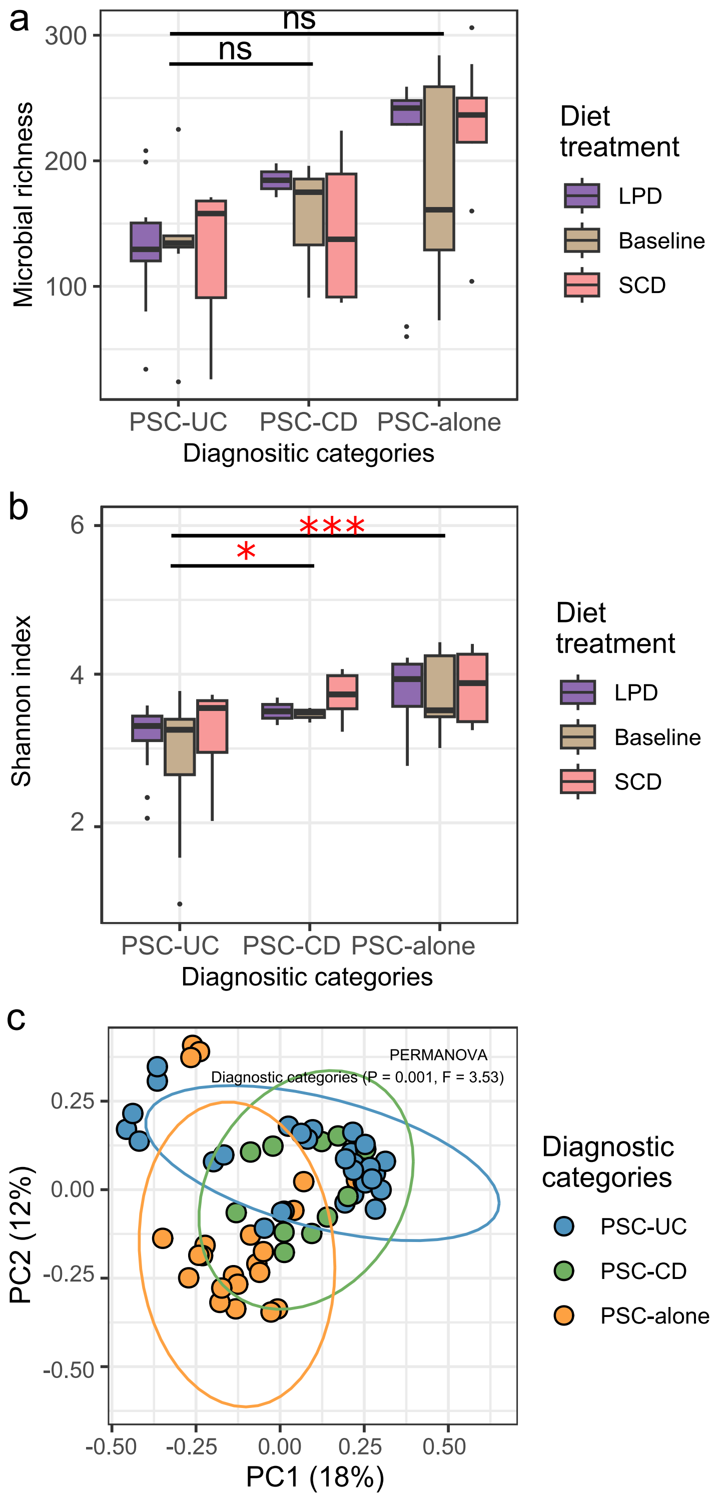


**Fig. S3.** Diagnostic categories (PSC-UC, PSC-CD, and PSC-alone) showed diverse microbiota, revealed by a) microbial richness (the number of detected species), b) Shannon index, and c) beta diversity. The Wilcoxon rank-sum test was employed to assess significance between disease categories, with *** denoting strong significance (p-value < 0.001), * denoting significance (0.01< p-value < 0.05), and 'ns' indicating no significant difference (p-value > 0.05).

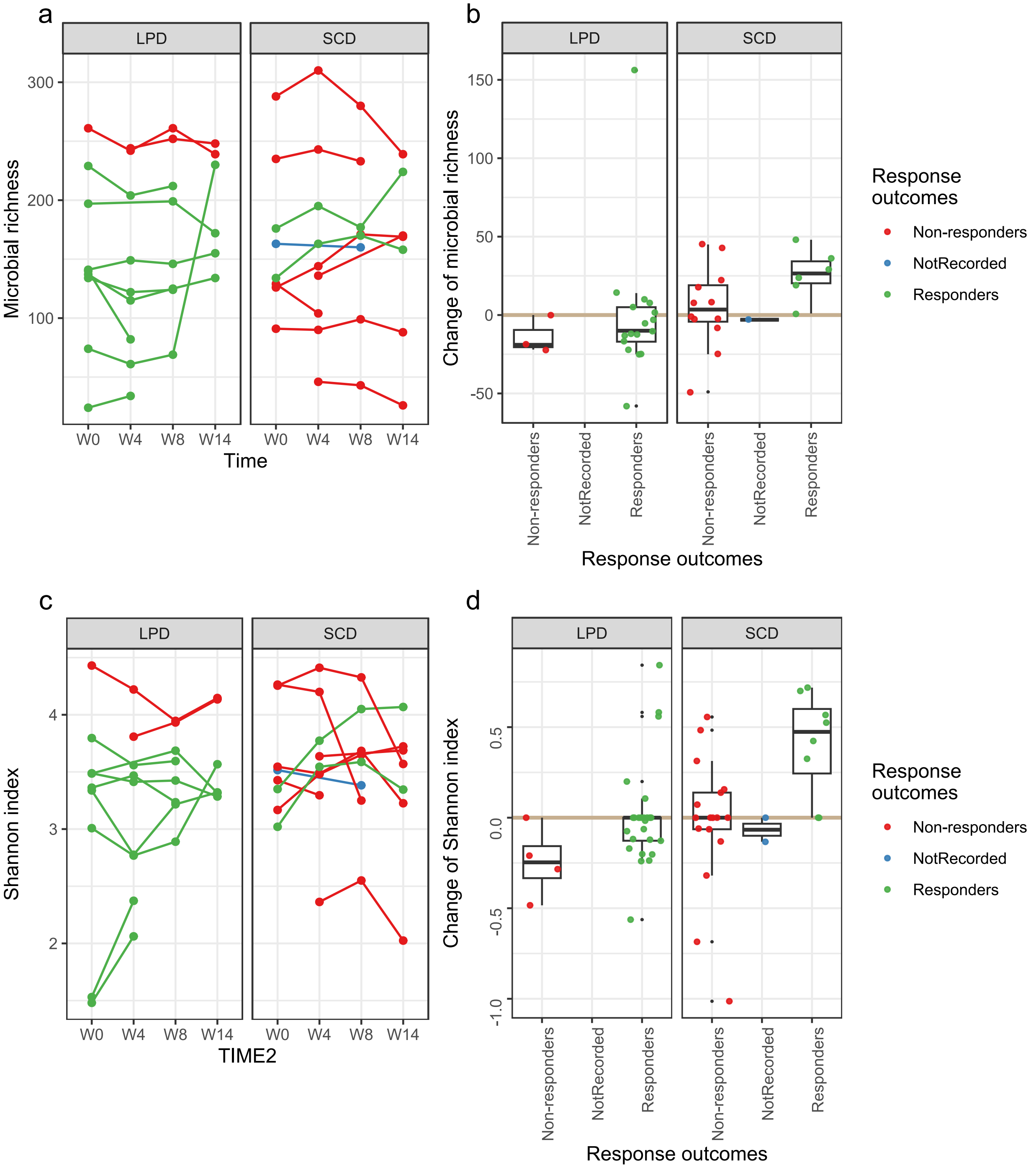


**Fig. S4.** Temporal variations of microbial alpha diversity revealed by the richness and Shannon index in each arm.

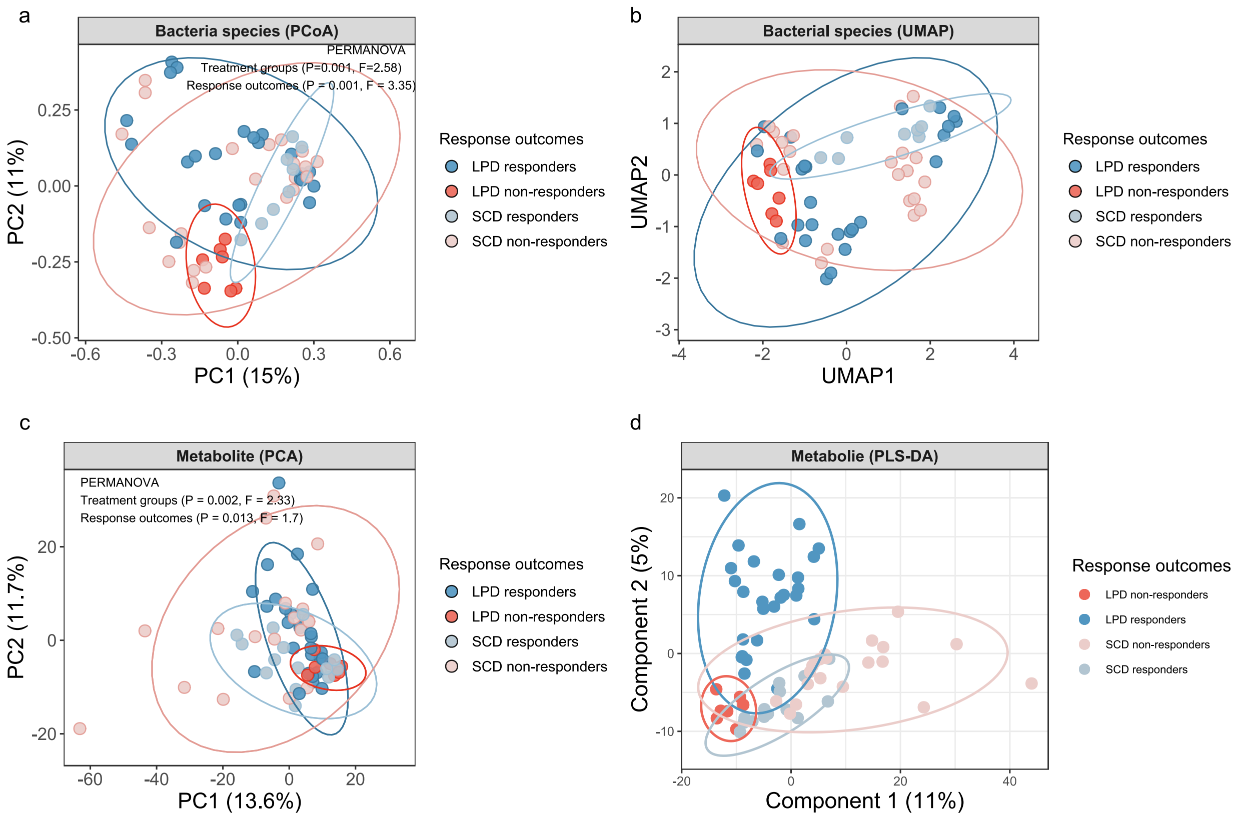


**Fig. S5**. Beta-diversity of bacterial species (a and b) and metabolites (c and d) across both arms (LPD and SCD).

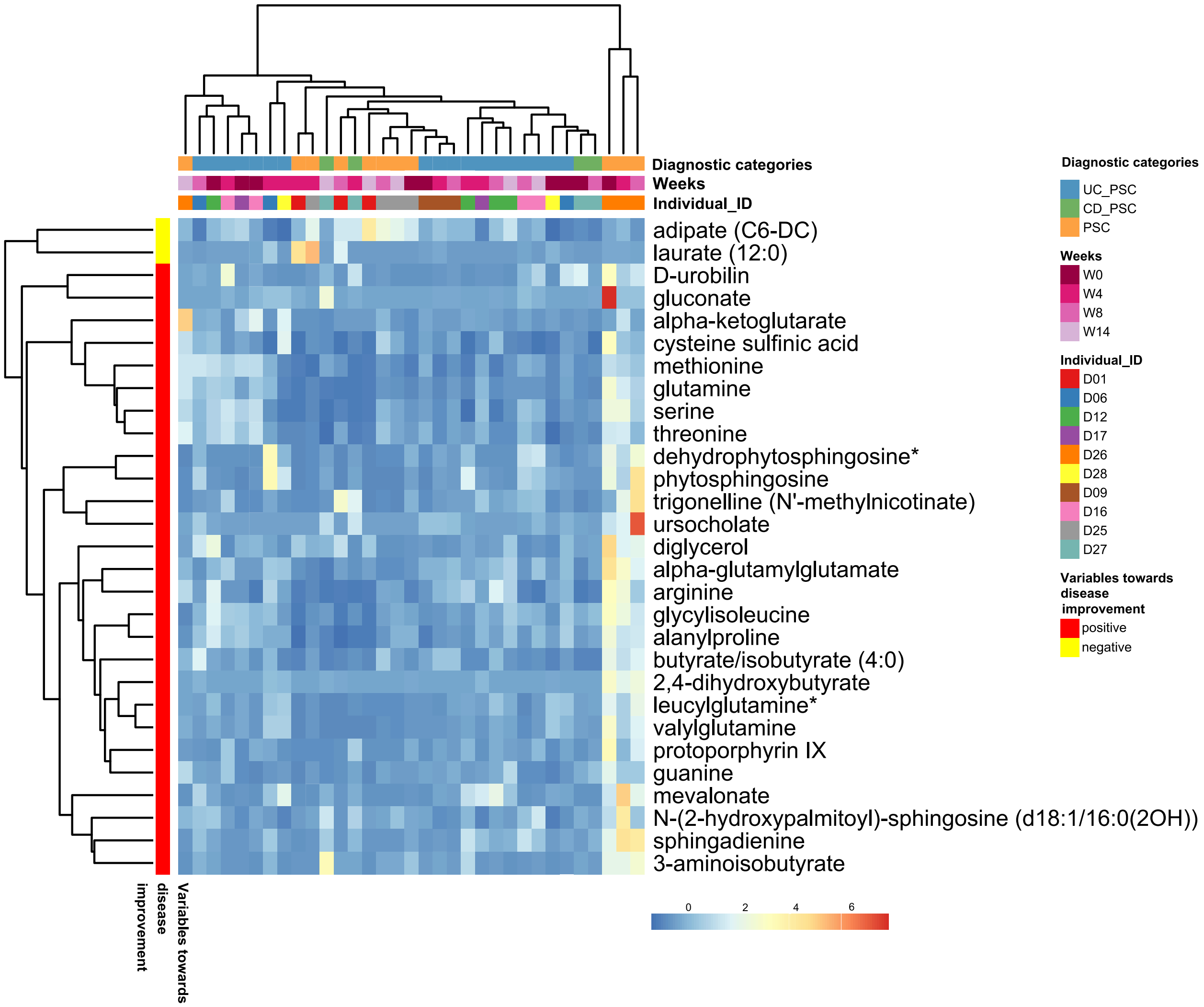


**Fig. S6.** Heatmap of metabolite markers that can distinguish LPD responders from non-responders, as determined by multivariate linear regression models (target p-value < 0.05 and FDR < 0.2). Similar to Fig. 3d, each column in the heatmap is a sample, and all the samples including both LPD responders and LPD non-responders are included. The only difference from Fig. 3d is that the clustering here is generated based on the similarity of the metabolite profiles in the samples, as opposed to the scheme in Fig. 3d, which uses direct clustering and column arrangement based on the order of the columns in the microbial profile heatmap.

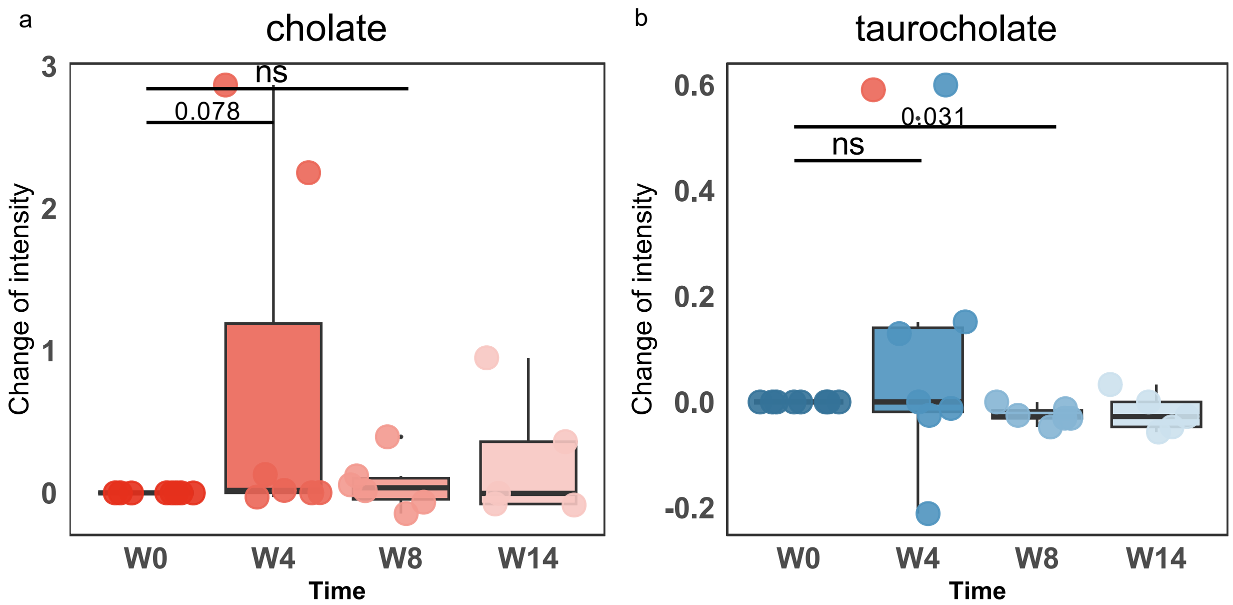


**Fig. S7.** Temporal changes in key bile acids, including cholate, and taurocholate (also named taurocholic acid). Bars are colored blue to represent metabolites that decreased after LPD, and orange for those that increased. The Wilcoxon signed-rank test was employed to assess significance between sampling time points, with ‘ns’ indicating no significant difference (p-value > 0.05).

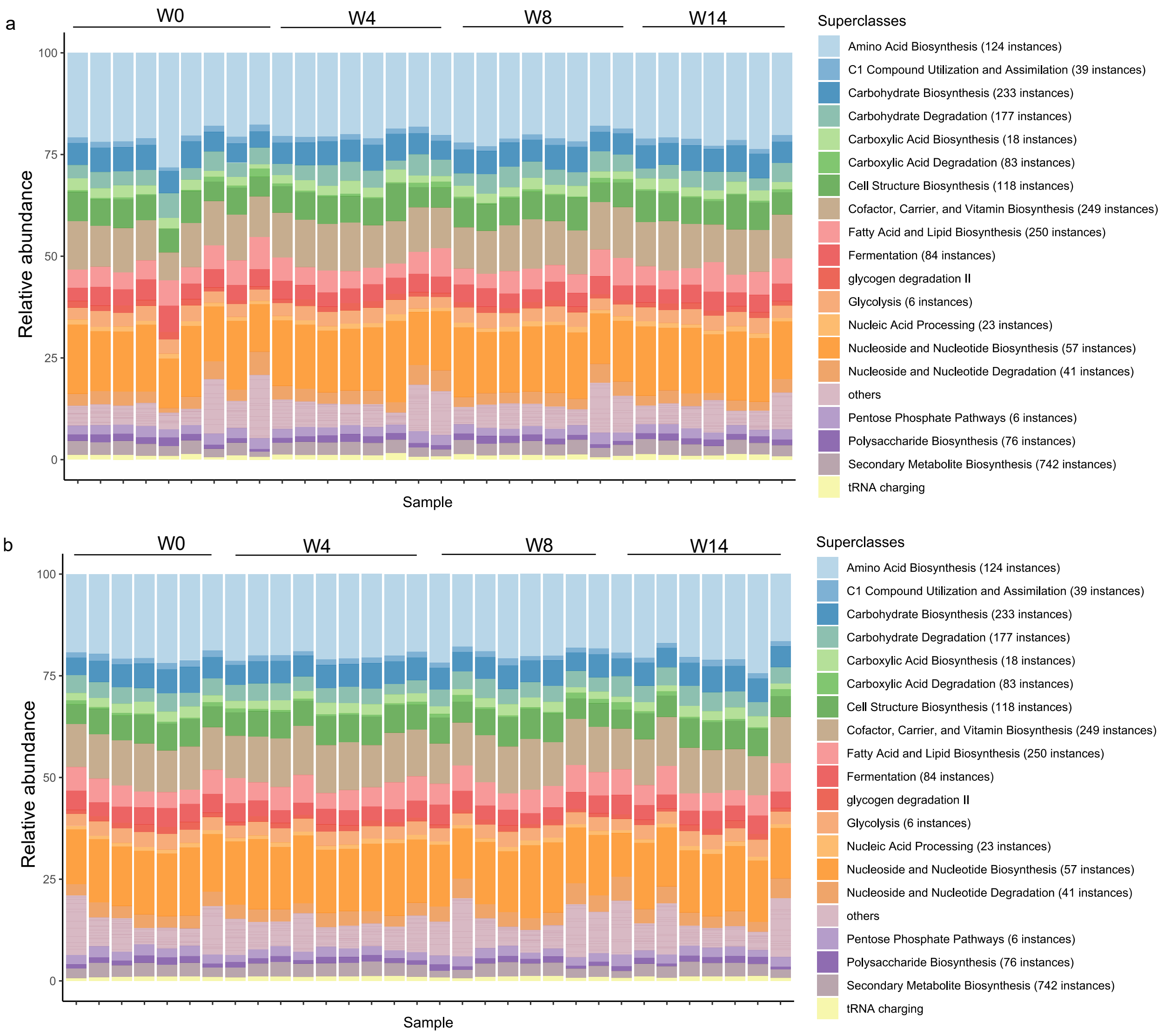


**Fig. S8.** Functional pathway classes across individuals and across time obtained from HUMAnN3.6 and referenced to the MetaCyc database. a) LPD and b) SCD are shown for two arms with different diet treatments. The most abundant 19 classes are shown, with the remainder categorized as 'Others'. W0, W4, W8, W14 represent samples from Week 0, Week 4, Week 8 and Week 14 respectively.

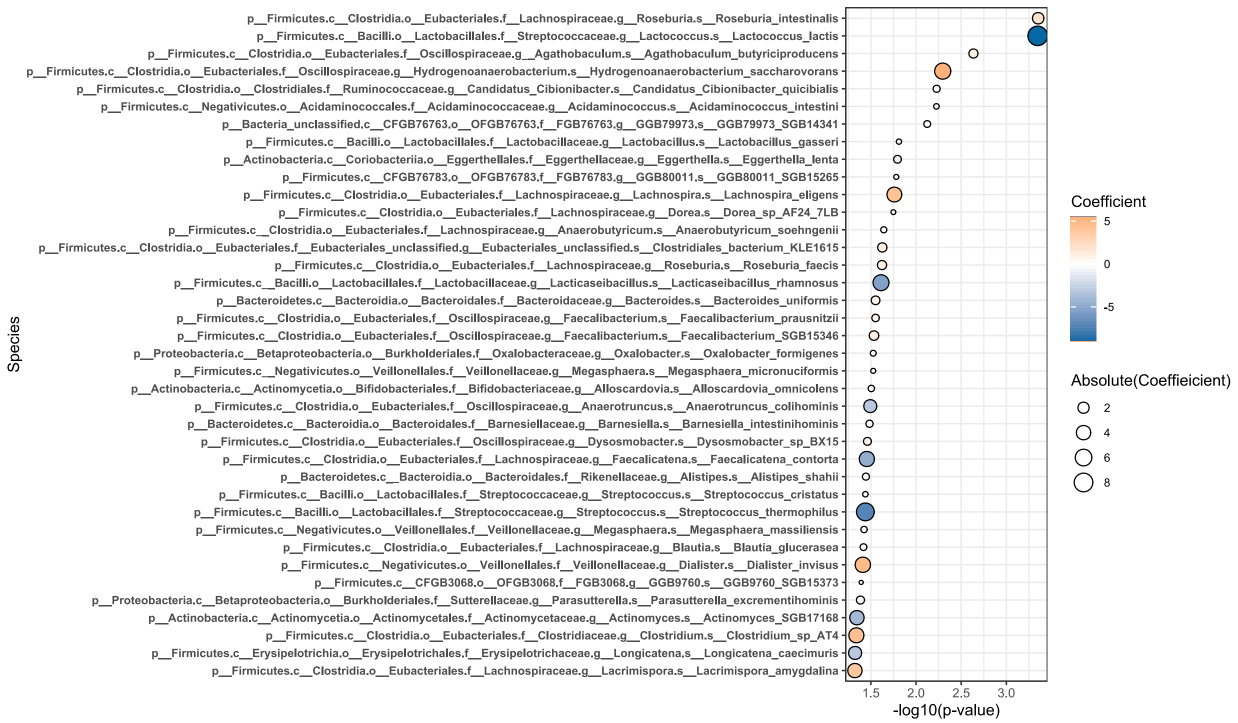


**Fig. S9.** Bacterial species exhibiting significant changes compared to baseline levels at Week 4 and Week 8 after LPD treatment, as revealed by a multivariate linear regression model (Formula 2, targeting p-values < 0.05). Species with a coefficient greater than 0 indicate an increase, while those less than 0 indicate a decrease.

**Table S6.** Detailed guidance for the Low Protein Diet (LPD) and Specific Carbohydrate Diet (SCD).

### Low Protein Diet

This is a specific diet that is both vegan and low in protein. The vegan diet eliminates all animal products, (including meats, eggs, dairy products) and animal by-products such as honey. Instead, all sources of protein are plant-based. This particular version of the vegan diet incorporates low-protein foods only, and aims for a maximum daily protein threshold of ____ (to be determined).

| | Allowed | NOT Allowed |
|---|---|---|
| FRUITS | • Any fresh/frozen fruit | • All dried fruits |
| VEGETABLES | • Any fresh/frozen/canned vegetables (except for those listed in "not allowed" column)<br>• Starchy vegetables: potatoes, yams, parsnips, winter squash, spaghetti squash, turnips, okra, bean sprouts<br>• Salads | • Spinach (boiled or raw), watercress, raw peas, raw mustard greens |
| PROTEIN | • Chestnuts, cashews, almonds, peanuts, brazil nuts, walnuts<br>• Coconuts<br>• Any beans: black beans, red kidney beans, chickpeas/garbanzo beans, mung beans, black eyed peas, etc.<br>• Lentils, seitan, chia seeds, flax seeds, hemp seeds, spirulina | • All meats: beef, lamb, pork, poultry, including processed meats (ex. hot dogs, bologna), smoked meats, and canned meats<br>• All fish and seafood (raw, cooked, or canned) |
| DAIRY | • None (only non-dairy/vegan products) | • Dairy milk of any kind (ex. yogurt, sour cream, buttermilk, half & half, butter, dairy chocolates) |
| GRAINS | • Wheat, corn, rye, rice, buckwheat, millet, triticale, bulgur, spelt, quinoa, rice (white/brown) | • Pearl barley, oatmeal, semolina, macaroni |
| BEVERAGES | • Any fresh juice: orange, grapefruit, grape, pineapple, pure apple cider, vegetable<br>• Herbal tea/coffee<br>• Non-dairy milk substitutes: coconut, hemp, rice, etc. (except for those listed in "not allowed column") | • Dairy milk |
| MISCELLANEOUS | • Salt, pepper, spices, dried or fresh herbs, lemon/lime juice, balsamic vinegar, olive oil, coconut oil, baking soda/baking powder, yeast<br>• Sugars and sweeteners: white/brown sugar, corn syrup, coconut sugar, raw cane sugar, any sugar substitute (ex. Splenda, Equal), maple syrup | • Soy-based products (tofu, soy sauce, soy-based meat alternatives, soy milk)<br>• Margarine, mustard, ground ginger |

Brigham and Women's Hospital
Contact: | (617) 732-7783

### Specific Carbohydrate Diet

The Specific Carbohydrate Diet emphasizes consumption of specific carbohydrates that require minimal digestion. Therefore, it eliminates most carbohydrates, including grains, starches, dairy and sugars. The following is a guide outlining which foods are allowed and which foods to avoid while on this diet.

| | Allowed | NOT Allowed |
|---|---|---|
| FRUITS | • Fresh, frozen, or dried fruit with no added sugar or other preservatives/additives | • Canned or dried fruit with added sugars or other additives |
| VEGETABLES | • Fresh or frozen non-starchy vegetables (with no added sugar or other preservatives/additives): see not allowed list, all others are allowed | • Potatoes, yams, parsnips, okra, turnips, bean sprouts, beans (soybean, mung bean, fava beans, chickpeas/garbanzo beans)<br>• Canned or packaged vegetables with additional sugars or preservatives |
| PROTEIN | • Almonds, pecans, brazil nuts, hazelnuts, walnuts, unroasted cashews, boiled chestnuts, peanut butter without any additives, other natural nut butters<br>• Fresh or frozen (with no added sugar or other preservatives/additives): Beef, lamb, pork, poultry, fish (including shellfish and canned fish in oil/water), eggs | • Shelled peanuts, nuts sold in salted mixtures<br>• Processed meats (ex. hot dogs, bologna), smoked meats (unless no sugar added in smoking process), canned meats, breaded fish, canned fish with sauces |
| DAIRY | • Natural cheeses (not ricotta or mozzarella), dry curd cottage cheese, home-made yogurt (see recipe in handout), lactose-free cheeses | • Processed cheeses, lactose-containing cheeses, buttermilk, sour cream, commercial yogurts |
| GRAINS | • NO GRAINS PERMITTED | • Wheat, barley, corn, rye, oats, rice, buckwheat, millet, triticale, bulgur, spelt |
| BEVERAGES | • Fresh juice with no sugar added: orange, grapefruit, grape, pineapple, pure apple cider, vegetable juices from any allowed vegetables, canned tomato juice<br>• Herbal tea/coffee<br>• Coconut and almond milk (homemade)<br>• Club soda<br>• Dry wine, gin, rye, scotch, bourbon, vodka | • Milk of any kind<br>• Coffee substitutes<br>• Soft drinks with fructose or glucose<br>• Beer, sweet wine, sherry, liqueurs, brandy |
| MISCELLANEOUS | • Butter<br>• Spices: dried or fresh herbs, lemon/lime juice, salt, pepper<br>• Sweeteners: honey, saccharine<br>• Baking soda | • Margarine<br>• Spice mixtures (ex. apple pie sauce, curry powder), garlic and onion powder<br>• Bouillon cubes or instant soup bases<br>• Ketchup<br>• Chocolate or carob, ice cream<br>• Sweeteners: molasses, corn syrup, maple syrup<br>• Baking powder |

Brigham and Women's Hospital
Contact: | (617) 732-7783
